# Weather, Social Distancing, and the Spread of COVID-19^*^

**DOI:** 10.1101/2020.07.23.20160911

**Authors:** Daniel J. Wilson

## Abstract

Using high-frequency panel data for U.S. counties, I estimate the full dynamic response of COVID-19 cases and deaths to exogenous movements in mobility and weather. I find several important results. First, holding mobility fixed, temperature is found to have a negative and significant effect on COVID-19 cases from 1 to 8 weeks ahead and on deaths from 2 to 8 weeks ahead. Second, holding weather fixed, mobility is found to have a large positive effect on subsequent growth in COVID-19 cases and deaths. The impact on cases becomes significant 3 to 4 weeks ahead and continues through 8 to 10 weeks ahead. The impact on deaths becomes significant around 4 weeks ahead and persists for at least 10 weeks. Third, I find that the deleterious effects of mobility on COVID-19 outcomes are far greater when the local virus transmission rate is above one – evidence supportive of public health policies aiming to reduce mobility specifically in places experiencing high transmission rates while relaxing restrictions elsewhere. Fourth, I find that the dynamic effects of mobility on cases are generally similar across counties, but the effects on deaths are higher for counties with older populations and, surprisingly, counties with lower black or hispanic population shares. Lastly, I find that while the marginal impact of mobility changes has been stable over recent weeks for cases, it has come down for deaths.

## 1 Introduction

Understanding the causal effects of social distancing behavior on subsequent growth in COVID-19 cases and deaths is clearly of first-order public health and economic importance. Public health officials need to know the impact of social distancing, which can be mediated by policy actions, on COVID-19 spread. In addition, individual mobility is integrally linked to economic activity.^1^ It is thus important for policymakers to understand the influence of policy-induced economic restrictions and reopenings on the spread of COVID-19, which in turn heavily affects the economic outlook going forward. As Jerome Powell, the Federal Reserve Chair, recently put it “Economic forecasts are uncertain in the best of times, and today the virus raises a new set of questions: How quickly and sustainably will it be brought under control? Can new outbreaks be avoided as social-distancing measures lapse?… The answers to these questions will go a long way toward setting the timing and pace of the economic recovery.”^2^ Yet, to date, there has been surprisingly little research estimating the full dynamic response of COVID-19 outcomes to exogenous movements in mobility.

There are a couple formidable empirical challenges that help explain this paucity of research. First, observed movements in mobility are likely to be endogenous due both to correlation with other factors that could themselves affect COVID-19 spread and to reverse causality, with publicity of current growth in cases and deaths affecting individuals’ mobility (voluntarily and/or via mandatory restrictions). Weather is one such factor. There have been numerous studies in recent months on the impact of temperature and other weather variables on COVID-19 spread. The results have been mixed, with some studies finding significant effects and some not. As this paper will show, weather and mobility are very strongly correlated and hence, given the possibility that weather has a direct effect on COVID-19 outcomes, it is important to study the impacts of weather and mobility *jointly*. Second, the potential lags between mobility and COVID-19 outcomes may be quite long, requiring at least several months of post-outbreak data before one can begin to trace out the full impacts of mobility changes. The availability of geographically granular, high-frequency, real-time data along with, of course, the passage of time now open the doors to such research.

This paper is a first attempt at estimating the full dynamic response of COVID-19 outcomes to exogenous movements in mobility. I estimate this impulse response function (IRF) for COVID-19 cases and deaths up to 10 weeks ahead using a panel Local Projections estimator with county-level data. To identify plausibly exogenous movements (“shocks”) in mobility, I use standard regression control techniques in a dynamic panel data framework.

In particular, when regressing future COVID-19 outcomes on current mobility, I control for lagged mobility, current and lagged COVID-19 outcomes, COVID-19 testing, weather (temperature, precipitation, and snowfall), and high-dimensional fixed effects for counties and for time. Controlling for current and lagged cases and/or deaths, as well as testing, accounts for the likelihood that news of current local COVID-19 spread, which itself would predict future cases and deaths, induces people to increase or decrease their current social distancing (mobility) behavior. Controlling for lagged mobility helps ensure that current movements in mobility are not driven simply by persistence from past mobility shocks. Including county fixed effects effectively controls for many important known and unknown characteristics of local communities that can increase COVID-19 transmission and/or lethality, such as demographics, socioeconomic status, density, etc. Time (week) fixed effects absorb seasonal factors, common time trends, and any policies or other factors at the national level. In an extension, I also show the results are robust to controlling for public health non-pharmaceutical interventions (NPIs), such as shelter-in-place orders and school closures.

The analysis reveals a number of important findings. First, holding mobility fixed, temperature is found to have a negative and significant effect on COVID-19 cases from 1 to 8 weeks ahead and on deaths from 2 to 8 weeks ahead, while precipitation and snowfall have no consistent significant effects. Second, holding weather fixed, mobility is found to have a large positive effect on subsequent growth in COVID-19 cases and deaths. The impact on cases becomes significant 3 to 4 weeks ahead and continues through 8 to 10 weeks ahead (depending on the measure of mobility). The impact on deaths becomes significant around 4 weeks ahead and persists for at least 10 weeks. The results are robust across a variety of mobility measures. Furthermore, the mobility effects are quantitatively large. For example, a 1% increase in time spent away from home is found to increase COVID-19 case growth over the following 8 weeks by about 1.8%.

Third, I find that the deleterious effects of mobility on COVID-19 outcomes are far greater when the local virus transmission rate is above one. When the transmission rate is below one, mobility has a small positive effect on subsequent cases but virtually no effect on deaths. When *R*_*t*_ is above one, mobility has a much larger effect on future cases and has a large, longlasting impact on future deaths. These results are consistent with epidemiological models that predict more infections for a given contact rate (which is closely linked to mobility) when the transmission rate is higher. They also provide empirical support justifying calls from some public health experts for mobility-reduction policies, such as shelter-in-place orders and restrictions on large gatherings, that specifically target places experiencing high transmission rates while not imposing such restrictions elsewhere.

Fourth, I find that the dynamic effects of mobility on cases are generally similar across counties, but the effects on deaths are higher for counties with older populations and, surprisingly, counties with lower black or hispanic population shares. Lastly, I find that while the marginal impact of mobility changes has been stable over recent weeks for cases, it has come down for deaths.

There are relatively few studies to date estimating the direct effects of mobility on COVID-19 outcomes. Prior studies have tended to focus on the effect of non-pharmaceutical policy interventions, especially shelter-in-place orders (“SIPOs”), on COVID-19 outcomes, rather than the direct effects of mobility.^3^ For instance, Courtemanche, Garuccio, Le, Pinkston, and Yelowitz (2020) finds a sizable impact of SIPOs on subsequent COVID-19 case growth in the U.S. Similarly, Hsiang, Allen, Annan-Phan, Bell, Bolliger, Chong, Druckenmiller, Huang, Hultgren, Krasovich, et al. (2020) exploit subnational panel data, including state-level data for the U.S., during the early months of the pandemic to estimate the impact of various NPIs on COVID-19 case growth, finding substantial beneficial impacts. These studies do not directly study the role of mobility, but rather assume that the effect of NPIs on COVID-19 cases is mediated through the channel of NPIs, reducing mobility which in turn reduces infections. Similarly, Askitas, Tatsiramos, and Verheyden (2020), which uses cross-country panel data in an event-study framework, finds that NPIs decrease the daily incidence of COVID-19. They also find that NPIs decrease mobility, though they do not assess whether NPIs affect COVID-19 cases independent of mobility or whether mobility directly affects cases.

There are a few other notable studies of the direct effects of mobility behavior on COVID-19 outcomes. Soucy, Sturrock, Berry, Westwood, Daneman, MacFadden, and Brown (2020) uses variation across 40 global cities for the late March to mid-April time period and find a strong correlation between mobility and COVID-19 case growth 14 days ahead. Badr, Du, Marshall, Dong, Squire, and Gardner (2020) also investigate the correlation between mobility and subsequent case growth, but using panel data from the 25 U.S. counties with the most cases as of mid-April. Estimating this correlation for varying lag lengths, they find it peaks at 11 days. These two studies are correlational. As noted above, the correlation between mobility and COVID-19 outcomes can be driven by omitted variables and/or reverse causality, such that correlations may be a misleading guide to the causal impacts of changes in mobility and thus less useful for policy prescriptions. Kapoor, Rho, Sangha, Sharma, Shenoy, and Xu (2020) is one of the first studies aiming to estimate the causal effect of mobility changes on COVID-19 cases. They investigate the early declines in mobility, prior to local NPIs, using a cross-sectional regression design with U.S. county-level data. They employ an instrumental variables approach, using precipitation as an instrument for the early mobility declines. They find that these declines were associated with fewer cases and deaths up to at least 18 days later, the farthest horizon they investigate. Their results are consistent with those in this paper, though I find effects persisting as far out as 10 weeks ahead. Most recently, Unwin, Mishra, Bradley, Gandy, Vollmer, Mellan, Coupland, Ainslie, Whittaker, Ish-Horowicz, et al. (2020) estimate the impact of mobility on the transmission rate, *R*_*t*_, of SARS-CoV-2, the virus that causes COVID-19. They estimate this impact with state panel data using a logit model with state-specific random effects. Similar to this paper, they measure mobility using the Google Mobility Reports. Because the true transmission rate is unknown, they infer it from observed COVID-19 deaths using a Bayesian semi-structural model. An important element of the model is an assumed lag structure between infection and death. The authors use estimates of that lag structure from early studies of the disease in China.

As mentioned above, the evidence on weather’s impact of COVID-19 is mixed. Xu, Rahmandad, Gupta, DiGennaro, Ghaffarzadegan, Amini, and Jalali (2020) finds a “modest” negative effect of temperature on covid19 case growth globally. They assume a 10-day lag, which the evidence in this paper suggests may be too short to see the full impact. Carleton, Cornetet, Huybers, Meng, and Proctor (2020) finds a negative relationship between UV light and COVID-19 case growth, while they find “weak or inconsistent lagged effects of local temperature, specific humidity, and precipitation.” Similarly, Jamil, Alam, Gojobori, and Duarte (2020) find no evidence of a link between temperature and COVID-19 case growth across countries and Chinese provinces. Yet, these papers do not generally account for the fact (documented below) that weather has very strong effects on mobility behavior, which itself affects COVID-19 outcomes, making it hard to know how much their results reflect a serious omitted variable bias.

Most of the studies above and others in the nascent literature on COVID-19 impose a specific response lag between mobility or NPI/SIPOs and COVID-19 cases. Typically, they assume lags of around 14 days, which is based on the logic of a roughly 7-day incubation period (from exposure to symptoms) and a 7-day “confirmation” period from symptom onset to a positive test result. Such an assumption may miss an important third phase, the transmission propagation phase. After that initial exposed individual becomes infectious, they may spread the infection to one or more additional individuals, who may in turn spread it to others, and so on. Each of these “rounds” of infection transmission involves its own incubation and confirmation delays, potentially spreading out over time the effect of any initial shock such as mobility, weather, NPIs, etc.. This highlights the usefulness of applying an estimation method, such as local projections, that does not impose any assumed response lag and instead allows for estimation of the full impulse response function.

## 2 Data and Stylized Facts

### 2.1 COVID-19 Data

Daily county-level on COVID-19 cases and deaths were obtained from usafacts.org, which compiled the data from state public health agencies.^4^ I also obtained alternative data on cases and deaths from the New York Times database (via tracktherecovery.org). The correlation between these two data sets is extremely high (0.9995) and the results in the paper are nearly identical using the New York Times data. Data on daily testing, which are only available at the state level, come from The COVID Tracking Project and were downloaded from tracktherecovery.org.

The national time series for COVID-19 cases and deaths are shown in Figure 1. The earliest cases occurred in late January, but nationally cases began accelerating rapidly in mid-March, with deaths accelerating around the beginning of April.^5^ Yet the upsurge in cases and deaths varied substantially across counties, as shown in the histogram in Figure 2. Of the 3,016 counties with available data on cases, about one-third had their first case in or before the third week of March (15th - 21st). The week with the most first cases, in 822 counties, was the following one, March 22-28. There is a long right tail thereafter. In fact, there remain a few dozen sparsely populated counties that have no cases as of **July 16**, the latest date of data as of this writing.

**Figure 1:**
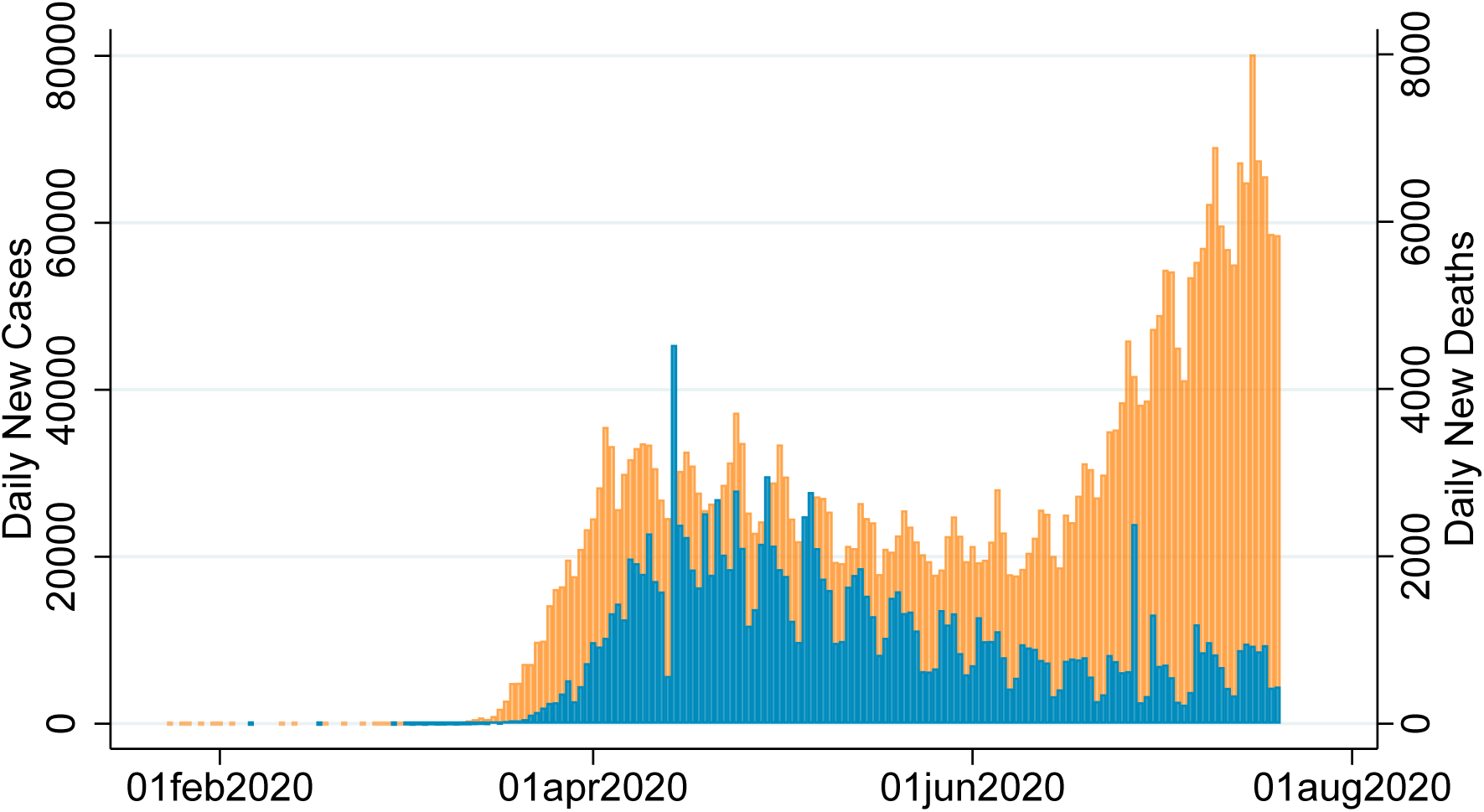
Daily COVID-19 Cases and Deaths in the United States

**Figure 2:**
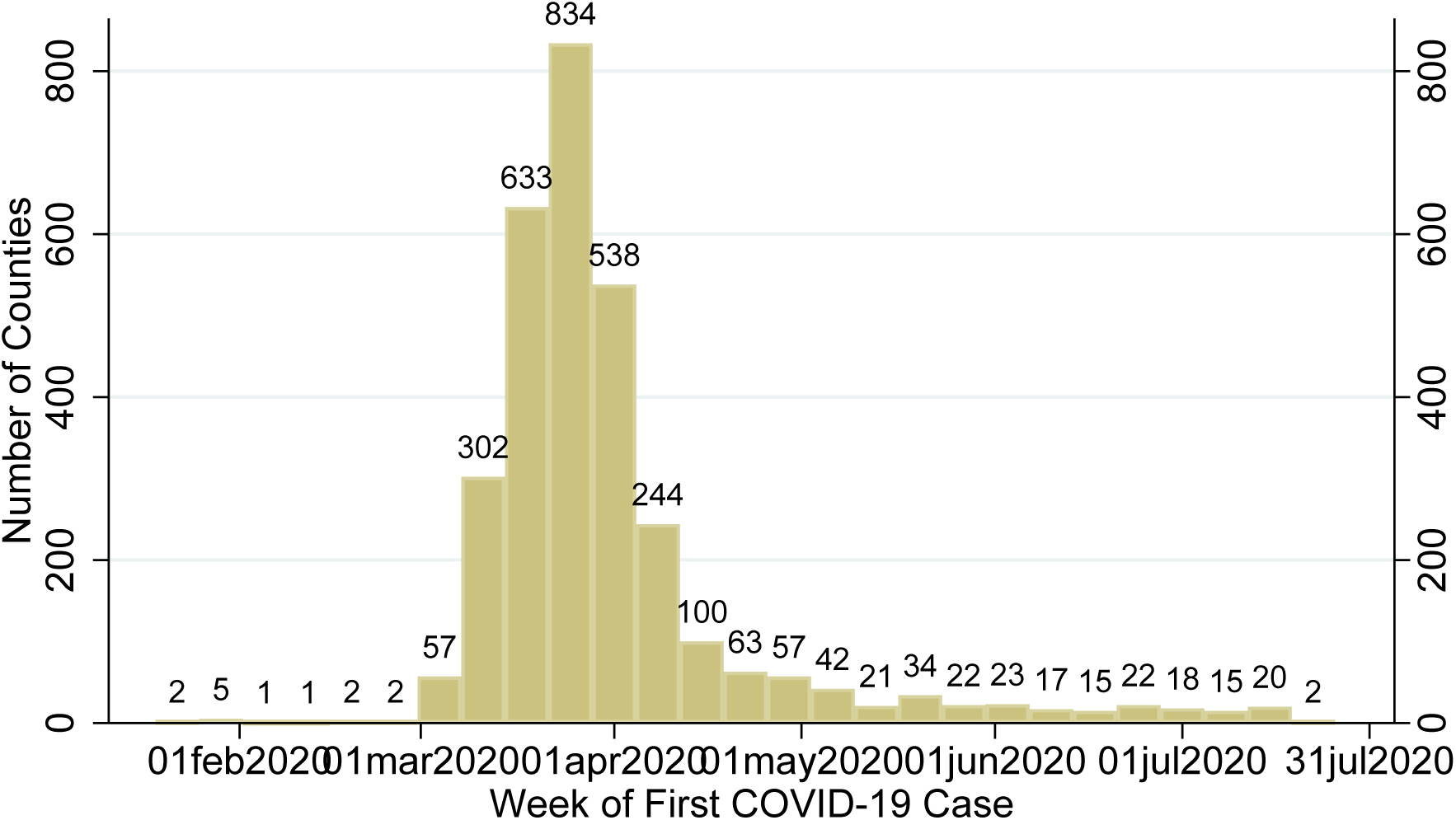
Timing of Onset of Local COVID-19 Outbreaks

### 2.2 Mobility Data

Google Mobility Reports provide percent changes in mobility relative to the Jan. 3 – Feb. 6 average. More specifically, Google describes the data as follows: “These datasets show how visits and length of stay at different places change compared to a baseline. We calculate these changes using the same kind of aggregated and anonymized data used to show popular times for places in Google Maps. Changes for each day are compared to a baseline value for that day of the week: The baseline is the median value, for the corresponding day of the week, during the 5-week period Jan. 3 – Feb. 6, 2020.”^6^ Google measures mobility separately for time spent at home, at workplaces, at transit stations, at grocery & pharmacy, and at retail & restaurants (which includes restaurants, cafes, shopping centers, theme parks, museums, libraries, and movie theaters). I multiply the time spent at home measure by −1 to facilitate comparison with the other measures. It should thus be interpreted as time spent *away* from home.

The Dallas Fed Mobility and Engagement Index (MEI) is the first principal component of seven cell-phone-based geolocation variables from the data provider Safegraph. The seven variables are as follows: the fraction of devices leaving home in a day, the fraction of devices away from home for 3-6 hours at a fixed location, the fraction of devices away from home longer than 6 hours at a fixed location, an adjusted average of daytime hours spent at home, the fraction of devices taking trips longer than 16 kilometers, the fraction of devices taking trips less than 2 kilometers, and the average time spent at locations far from home. Each variable is scaled by the weekday-specific average over January and February prior to the principal component analysis. See Atkinson, Dolmas, Koch, Koenig, Mertens, Murphy, and Yi (2020) for details.^7^

Figure 3 shows how these mobility measures have evolved since late January nationally as well as in some selected cities. For the nation as a whole, mobility plunged over the second half of March, bottomed out in early April, and gradually recovered thereafter, though it has stalled somewhat in July. Yet, there is considerable variation across localities in the patterns of mobility. Some places, such as Seattle (King County, WA), San Francisco (San Francisco County, CA), and New York City (counties of New York, Kings, Queens, Bronx, and Richmond), saw mobility fall much sooner and further than it did nationally. In other places, such as Phoenix (Maricopa County, AZ), mobility fell around the same time as it did nationally but by a smaller amount. The recovery pattern has also varied substantially across places. For example, mobility in New York City recovered relatively quickly and steadily since mid-April, while mobility in Phoenix rose slowly from mid-April to early June and has fallen slightly since then. It is this type of variation – that is, the variation across counties in their differences from the national average – that I exploit to estimate the impact of mobility on subsequent COVID-19 outcomes.

**Figure 3:**
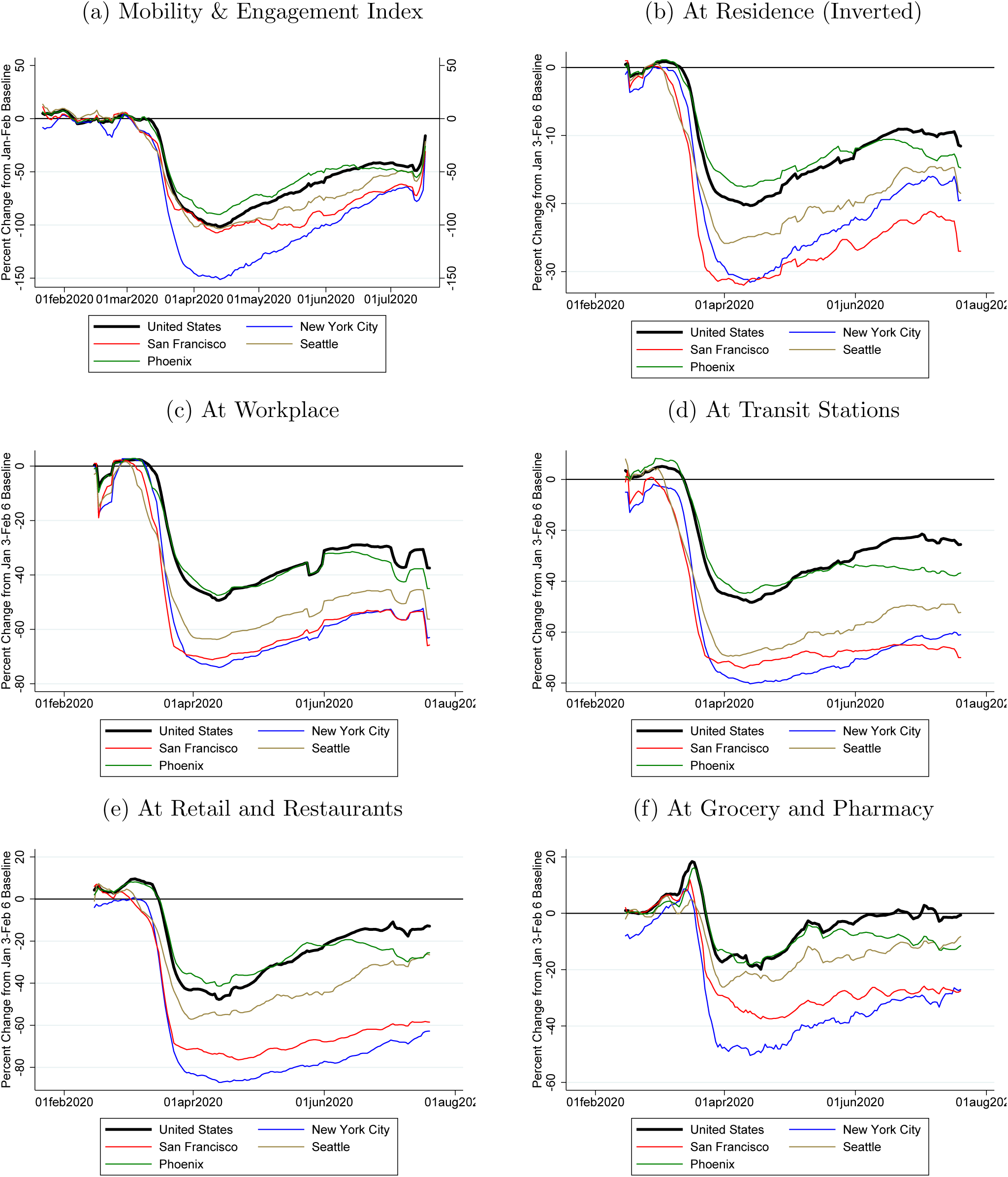
Mobility Over Time

### 2.3 Weather Data

Following Wilson (2019), I construct measures of daily weather at the county level from the Global Historical Climatology Network Daily (GHCN-Daily) data set. The GHCN-Daily is provided by the U.S. National Climatic Data Center (part of the National Oceanic and Atmospheric Administration (NOAA)) and contains daily weather measurements from a little over 4,700 weather stations throughout the United States, though not all stations provide readings every day. All stations with data on a given date are used for measuring county weather on that date. The spatial distribution of weather stations is highly correlated with the spatial distribution of population.

The readings from individual weather stations are used to estimate county-level weather using an inverse-distance weighting procedure. First, the surface of the conterminous United States is divided into a 5-mile by 5-mile grid. Second, weather values for each grid point are estimated using inverse-distance-weighted averages of the weather values from weather stations within 50 miles of the grid point. See Wilson (2019) for further details of this procedure.

This procedure yields the following daily county-level weather variables: maximum temperature (degrees Fahrenheit), minimum temperature, precipitation (mm), and snowfall (cm).

### 2.4 Other Data

I obtained data on start- and end-dates of various local non-pharmaceutical interventions (NPIs) from the Keystone *Coronavirus City And County Non-Pharmaceutical Intervention Rollout Date Dataset*.^8^ Keystone has compiled data for all states and about 600 counties, including all counties that had at least 100 cases as of April 6, 2020. I use the county level data for those ≈600 counties and state level data for other counties. The data include start- and end-dates for the following 10 NPIs: social distancing (“social distancing mandate of at least 6 feet between people”), shelter in place (“an order indicating that people should shelter in their homes except for essential reasons”), prohibition of gatherings above 100, prohibition of gatherings above 25, prohibition of gatherings above 10, prohibitions of gatherings of any size, closing of public venues (“a government order closing gathering venues for in-person service, such as restaurants, bars, and theaters”), closure of schools and universities, closure of non-essential services and shops, closure of religious gatherings, and full lockdown. I condense these data into a single variable, the number of NPIs in place in the county on each date.

Data on population (average daily residents) in nursing homes by county as of June 4, 2020 were obtained from the Centers for Medicaid and Medicare at: https://data.medicare.gov/Nursing-Home-Compare/Provider-Info/4pq5-n9py.

## 3 Methodology

My primary objective is to estimate the causal effects of mobility, which can be influenced by government policy choices and public opinion, on COVID-19 spread. Simultaneously estimating the causal effects of weather, which is exogenous, on COVID-19 is both a secondary objective and necessary for obtaining an unbiased estimate of mobility’s effect given that weather and mobility are likely correlated. Below I describe the econometric methodology used to achieve these objectives.

### 3.1 Contemporaneous Effect of Weather on Mobility

I first estimate the contemporaneous effect of weather (**w**) on mobility (m) using a daily county panel data model:

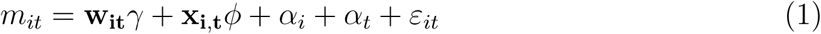

where *i* indexes counties and *t* indexes dates. **w**_**it**_ is a vector of weather variables, consisting of daily maximum temperature, precipitation, and snowfall. **x**_**i**,**t**_ is a vector of control variables, consisting of COVID-19 case growth (daily new cases divided by total cases) and testing growth. *α*_*i*_ and *α*_*t*_ are fixed effects for county and date. The rationale for the inclusion of these control variables and fixed effects is discussed in the *Identification* subsection (3.3) below.

### 3.2 Dynamic Effects of Mobility and Weather on COVID-19

Next, I jointly estimate the effects of mobility (*m*) and weather (**w**) on subsequent COVID-19 growth (*g*) using a panel Local Projections estimator:

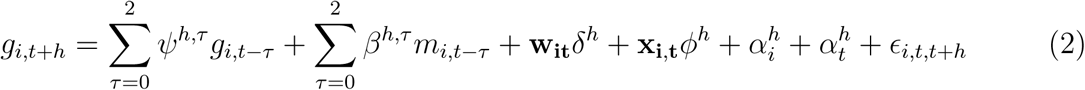

where *i* indexes counties, *t* indexes time (days for daily regressions and weeks for weekly regressions), and **x**_**i**,**t**_ is a vector of control variables.

I analyze two COVID-19 outcomes (*g*): growth in cases and growth in deaths. Growth during a given time period (*t* + *h*) is defined as the number of new cases (deaths) recorded during that period divided by the total number of cases (deaths) as of time *t*. For instance, case growth during period *t* + *h* is calculated as *g*_*i,t*+*h*_ = (*cases*_*i,t*+*h*_ − *cases*_*i,t*+*h*−1_)*/cases*_*i,t*_. Note that this variable is the *flow* of new cases (deaths) during *t* + *h*.

To additionally estimate effects on *cumulative* growth of cases or deaths over a given horizon, I estimate versions of equation 2 in which the dependent variable is cumulative growth (*G*), defined as the cumulative sum of new cases from *t* to *t* + *h* divided by the total number of cases (deaths) as of time *t*: *G*_*i,t*+*h*_ = (*cases*_*i,t*+*h*_ − *cases*_*i,t*_)*/cases*_*i,t*_, and similarly for deaths.

The local projections method (Jord`a (2005)) traces out an impulse response function (IRF) by estimating equation 2 sequentially over horizons from *h* = 1 to some maximum horizon, *H*. I estimate IRFs out to *H* = 10 weeks ahead. The IRF for mobility is traced out by the sequence of *β*^*h*,0^, while the IRF for any element of the weather vector **w**_**it**_ is traced out by the sequence of its element of the coefficient vector *δ*^*h*^.

I estimate IRFs at a weekly frequency, using weekly aggregated data, for three reasons. First, aggregating the daily data to weekly frequency removes the sizable variation between weekdays and weekends in the time series of mobility as well as COVID-19 cases and deaths. Second, there likely is considerable measurement error at the daily frequency in COVID-19 cases, deaths, and testing due to reporting lags. That measurement error should largely cancel out with aggregation to the weekly level. Third, estimating equation 2 is computationally intensive and hence estimating it for daily horizons from *h* = 7 to 70 would be extremely time intensive. Nonetheless, for robustness, I also produce IRF results at the daily frequency for horizons 7, 14,…,70. The results are very similar to the weekly results, though less precisely estimated.

### 3.3 Identification

The causal effect of mobility, *β*^*h*,0^, or weather, *δ*^*h*^, on COVID-19 cases and deaths is unlikely to be identified by any simple cross-sectional correlations due to a variety of omitted variable and reverse causality concerns. I address these identification concerns through dynamics, control variables, and fixed effects. In terms of dynamics, equation 2 uses leads of the COVID-19 outcomes as dependent variables to mitigate the potential contemporaneous reverse causality due to local news about current cases or deaths inducing people to increase or decrease their social distancing (mobility) behavior. For the same reason, I include the contemporaneous value and two weekly lags of the dependent variable as well as current growth in testing. Controlling for current testing helps mitigate concerns that public fears (or lack thereof) about community spread may affect mobility and also be correlated with future cases and deaths growth because such fears should be reflected in greater demand for tests.

When the dependent variable is growth in deaths, I also add current case growth as a regressor. I additionally include two weekly lags of mobility so that *β*^*h*,0^ can be interpreted as the effect of a current “shock” or change in mobility that was not driven simply by its recent trend.

As the results below will demonstrate, when studying the effect of mobility on COVID-19, it is important to control for weather. Likewise, when studying the effect of weather on COVID-19, it is important to control for mobility. As discussed in the introduction, most COVID-19 studies to date of either weather or mobility have not controlled for the other factor and could be subject to serious omitted variable bias. Thus, I include both mobility and weather variables – maximum daily temperature, precipitation, and snowfall – in all regressions unless otherwise indicated.

The county fixed effects absorb many important known and unknown characteristics of local communities that can increase COVID-19 transmission and/or lethality. These time-invariant characteristics include demographics (such as age, gender, and race), socioeconomic status, access to healthcare, population density, average household size, the presence of nursing homes or meat-packing plants, and openness to international travelers. Desmet and Wacziarg (2020) document the importance of many such time-invariant factors for COVID-19 cases and deaths among U.S. counties. In Section 5.1, I investigate whether the average “treatment” effect of mobility on COVID-19 varies across some of these county characteristics.

The time fixed effects are also crucially important. In particular, they will absorb seasonal factors and any common (i.e., national) time trends. This is particularly important given that weather, especially temperature and snowfall, obviously has strong trends over the January to June sample period, and mobility has also trended higher from late March onward.

### 3.4 Inference

The standard errors and confidence intervals reported in the paper are robust to heteroskedasticity and two-way clustering by county and state-by-time (where time is date for daily regressions and week for weekly regressions). The clustering by county allows for the possibility that errors, *ε*_*i,t*_ in equation 1 and *ϵ*_*i,t,t*+*h*_ in equation 2, are serially correlated. The clustering by state-time allows for the possiblity that errors are contemporaneously correlated across counties within the same state. This clustering will account for cross-county correlation stemming from unobserved statewide factors such as state economic and public health policies. It will also account for geo-spatial correlation in measurement error, for example in weather data, to the extent such correlation is encompassed by state boundaries.

### 3.5 Data Sample

All regressions in the paper use the maximum data sample available for the variables used in that regression, with one restriction. I restrict the sample time period for each county to begin with the first date on which cases per capita exceeded one per 1,000 persons. This restriction excludes observations from time and places where the COVID-19 outbreak had not yet begun. The sample time period varies across regressions depending on the horizon (in the local projections regressions) and on the data availability of the mobility variable used. For the local projections regressions, the further out the horizon, the fewer the time periods (*t*) available for estimation.

Data availability varies across the mobility variables, with the MEI data beginning in January and the Google Mobility data beginning in mid-February. All variables are available through late June as of the time of this writing. None of the mobility variables is available for all counties due to suppression of data (by Safegraph and Google) for counties with fewer mobile devices to mitigate privacy concerns. The county coverage varies across the Google mobility measures from about 1,100 counties for time spent at transit stations to roughly 2,800 for time spent at work. The MEI data covers about 3,000 counties. (There are 3,140 counties in the U.S..)

## 4 Main Results

### 4.1 Effects of Weather on Mobility

Before presenting the formal regression results, I begin with some non-parametric graphical evidence on the contemporaneous daily relationship between temperature and mobility. Temperature is the maximum daily high measured in degrees Fahrenheit. The panels in Figure 4 show bin-scatter plots with temperature on the x-axis and mobility on the y-axis. A bin-scatter plot is a common way to visualize correlations involving a large number of observations. Each variable is first residualized by regressing on county and date fixed effects. The × and y variables are then averaged within 100 bins corresponding to each percentile of the distribution of temperature values. (Hence, bins will be many degrees wide toward the lower and upper end of the temperature range and somewhat narrower than one degree in the middle of the range.) These plots show a strong positive relationship between temperature and mobility, conditional on county and time fixed effects. For the Google Mobility measures, the relationship is approximately linear, though some measures show a flat relationship as temperature rises above around 80*°* F. This flattening at high temperatures is more apparent for the MEI and starts around 70*°* F. The tightness of the fit varies across mobility measures. It is tightest for time spent at home, retail & restaurants, and at grocery & pharmacy.

**Figure 4:**
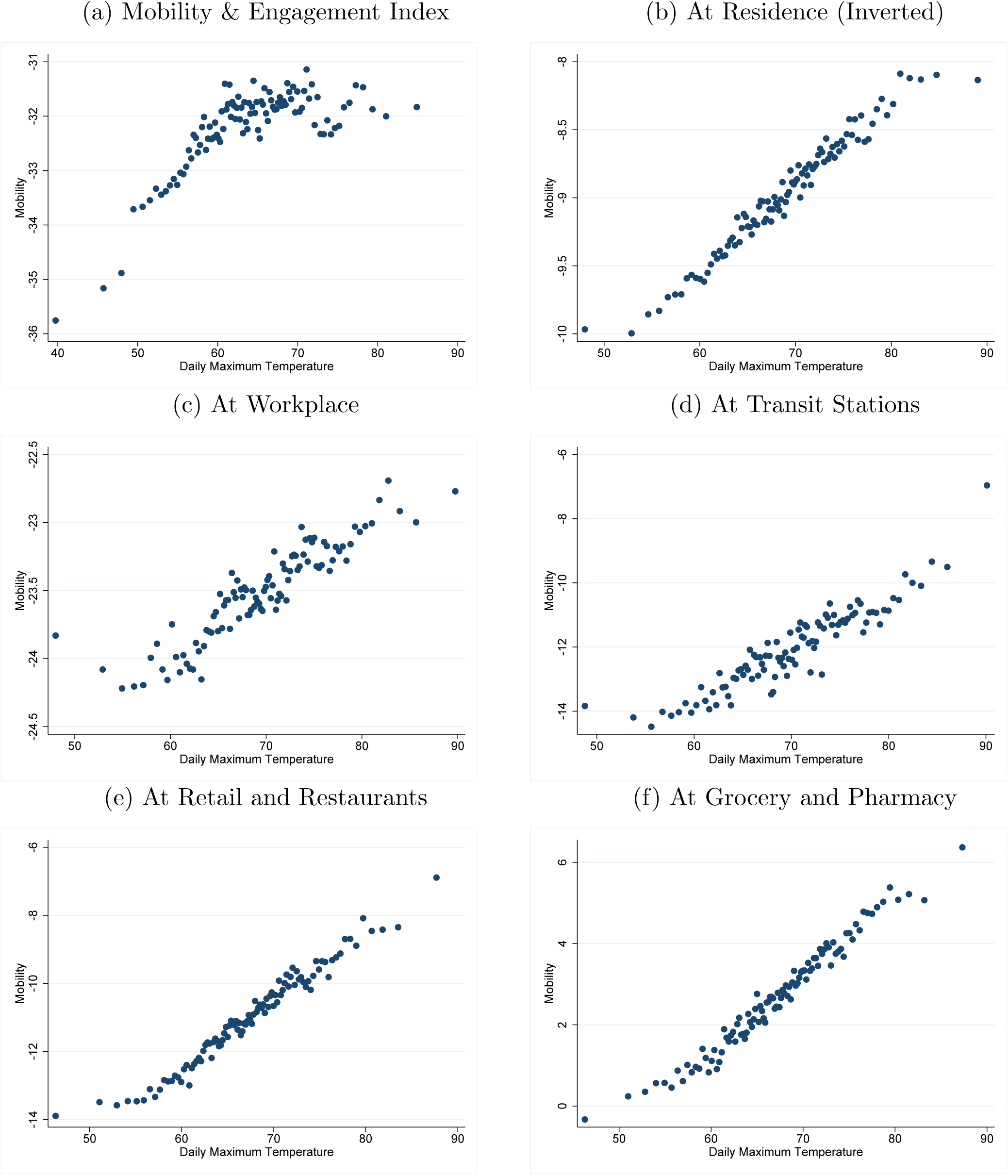
Relationship Between Temperature and Mobility Note: Bin scatterplots, using 100 bins after residualizing on county and date fixed effects.

Table 1 displays the results from estimating equation 1 using alternative measures of mobility. Each column corresponds to a separate regression and the column heading indicates which mobility measure is used as the dependent variable. The mobility measures are the six Google Mobility Report variables (“At…”) and the Dallas Fed Mobility & Engagement Index (MEI). The sample time period and number of counties are indicated at the bottom of the table and vary primarily depending on the availability of the mobility data, as noted in the previous section. Recall that county*date observations in which cases did not yet exceed one per 10,000 population are excluded, thus each regression uses an unbalanced panel. The weather variables are interacted with a weekday vs weekend indicator to allow for the possiblity that weather affects mobility differently on weekdays, when most workers work, than on weekends, when most workers have off.

**Table 1:** Effect of Weather on Mobility

|  | (1)<br>MEI | (2)<br>At Home (Inverted) | (3)<br>At Work | (4)<br>At Transit | (5)<br>At Retail & Rest. | (6)<br>At Grocery & Pharmacy |
| --- | --- | --- | --- | --- | --- | --- |
| Max Daily Temp – Weekday | 0.0568***<br>(0.0133) | 0.0699***<br>(0.00316) | 0.0614***<br>(0.00594) | 0.202***<br>(0.0182) | 0.213***<br>(0.0133) | 0.189***<br>(0.0116) |
| Max Daily Temp – Weekend | -0.0411**<br>(0.0175) | -0.000169<br>(0.00521) | -0.0298***<br>(0.00877) | 0.0269<br>(0.0238) | 0.120***<br>(0.0167) | 0.140***<br>(0.0152) |
| Precipitation – Weekday | -0.147***<br>(0.0117) | -0.0221***<br>(0.00287) | -0.0117***<br>(0.00404) | -0.0399***<br>(0.00892) | -0.0346***<br>(0.00980) | -0.0324***<br>(0.00872) |
| Precipitation – Weekend | -0.164***<br>(0.0212) | -0.0338***<br>(0.00703) | -0.0243**<br>(0.0106) | -0.0896***<br>(0.0219) | -0.0699***<br>(0.0196) | -0.0875***<br>(0.0203) |
| Snowfall – Weekday | -0.777***<br>(0.159) | -0.103***<br>(0.0358) | -0.221***<br>(0.0608) | -0.201*<br>(0.116) | -0.405***<br>(0.0935) | -0.421***<br>(0.0829) |
| Snowfall – Weekend | -0.469***<br>(0.173) | -0.278***<br>(0.0582) | -0.298***<br>(0.0942) | -1.000***<br>(0.272) | -0.508***<br>(0.139) | -0.419***<br>(0.144) |
| Observations | 465838 | 164479 | 346548 | 135915 | 249117 | 231964 |
| Adjusted $R^2$ | 0.858 | 0.910 | 0.892 | 0.745 | 0.799 | 0.660 |
| Earliest date | 22jan2020 | 15feb2020 | 15feb2020 | 15feb2020 | 15feb2020 | 15feb2020 |
| Latest date | 11jul2020 | 15jul2020 | 15jul2020 | 15jul2020 | 15jul2020 | 15jul2020 |
| # of days | 171 | 152 | 152 | 152 | 152 | 152 |
| # of counties | 2823 | 1485 | 2594 | 1080 | 2399 | 2299 |

\* $p < 0.10$ , \*\* $p < 0.05$ , \*\*\* $p < 0.01$

The results show a number of clear patterns. First, weekday temperatures have a strong positive effect on all measures of mobility. The effect is strongest for time spent at retail and restaurants. The coefficient of **0.213** (standard error of **0.013**) implies that 10 degrees of higher temperature on a weekday increases mobility at retail and restaurants by a little over **2** percentage points. (Recall that the Google mobility variables are measured in percentage change relative to Jan. 3 – Feb. 6.) Second, weekend temperatures have a positive effect on some mobility measures (retail & restaurants and grocery & pharmacy) but a negative effect on others (work and MEI). The negative effect may reflect that some workers with flexibility regarding weekend work may opt to choose leisure over work on weekends with pleasant weather (and/or on hot days when outside work is less pleasant). Third, precipitation has a strong negative effect on all measures of mobility. The negative effect is true for precipitation on both weekdays and weekends, though it larger for weekends. This likely reflects that common weekend activities, like going out to eat, going retail shopping, and going grocery shopping are less appealing when it is raining. Finally, snowfall, like rain, also has a strong negative effect on mobility, with larger effects on weekends.

Overall, Table 1 makes clear that weather and mobility are highly correlated and thus omitting either factor when studying the COVID-19 effects of the other is likely to result in an omitted variable bias.

### 4.2 Dynamic Impacts of Weather on COVID-19

I now turn to jointly estimating the impulse response functions (IRFs) of mobility and weather on COVID-19 cases and deaths using the local projections estimator described above (equation 2). As mentioned earlier, I estimate the IRFs at the weekly frequency on weekly-aggregated data in order to smooth over the day-of-week variability and high-frequency measurement error and also to reduce computational burden. The dependent variables are *h*-weeks ahead growth in cases and deaths, for *h* = 1 to 10 weeks. Recall that growth is defined as new cases (deaths) during week *t* + *h* relative to total cases (deaths) as of week *t*. I begin by discussing the estimated IRFs with respect to weather shocks. The results are presented in Figure 5. The IRFs on the left column are for growth in cases, while those on the right are for growth in deaths. The point estimates are shown with the circles, while the inner and outer brackets display the 90 and 95% confidence intervals, respectively. The regressions underlying Figure 5 use the Google Mobility measure of time spent away from home, though the results are similar using the other broad mobility measure, the MEI.

**Figure 5:**
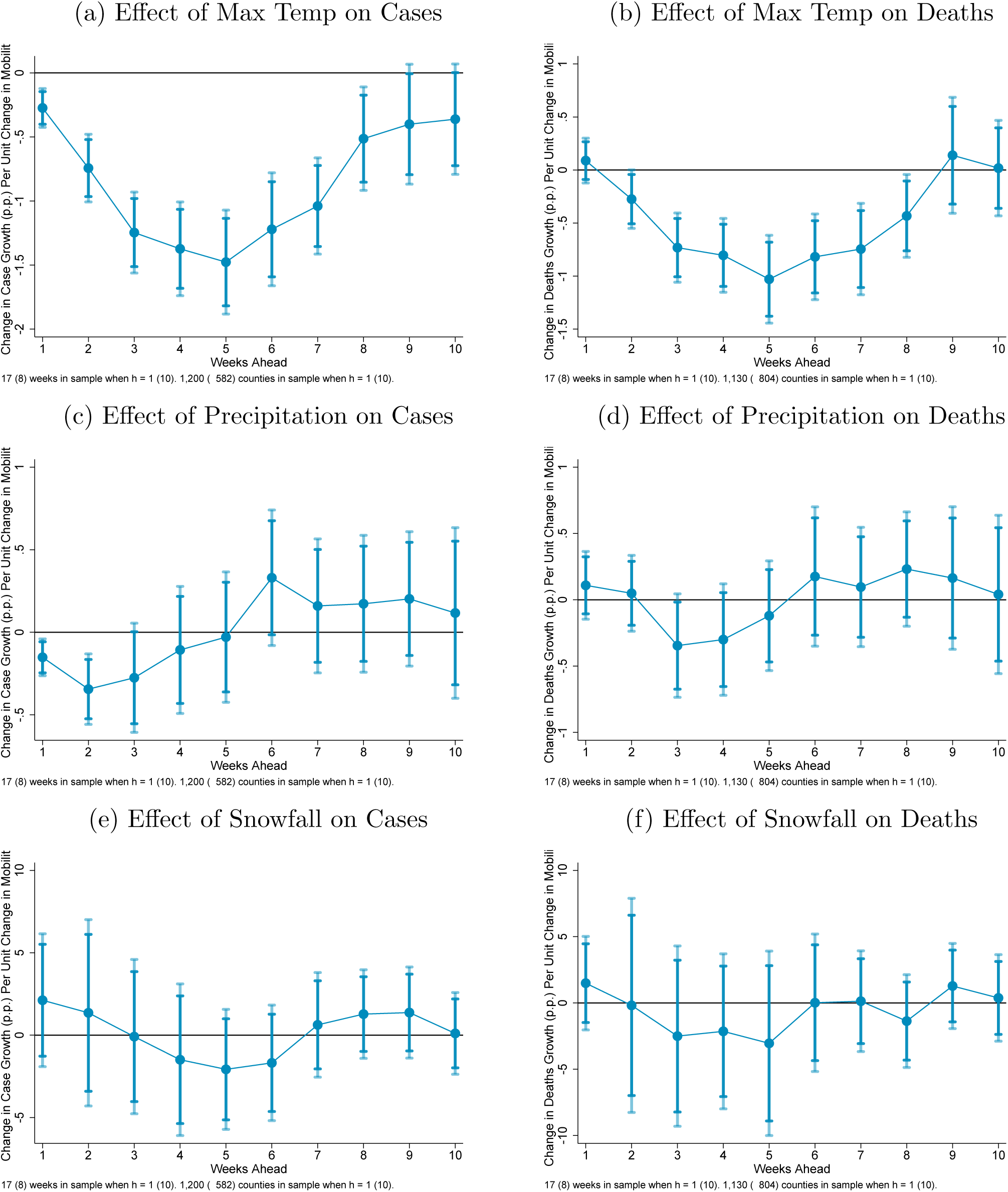
Dynamic Impacts of Weather on COVID-19 Case Growth Impulse Response Functions Estimated by Panel Linear Projections Note: Estimates of equation 2 in the text using panel local projections regressions. Shaded regions are 90% and 95% confidence intervals.

Holding mobility fixed, temperature is found to have a large and long-lasting negative effect on COVID-19 case growth. Specifically, higher temperatures reduce case growth starting as soon as 1 week ahead and for up to 8 weeks ahead. The effects after 8 weeks are only marginally statistically significant. The peak coefficient on temperature is approximately **-1.5** at 5 weeks ahead, with a 95% confidence interval of **-1.1** to **-1.9**. This coefficient implies that one degree higher daily maximum temperature during a week lowers growth in COVID-19 cases 5 weeks later by **1.5** percentage points. Evaluated at the sample mean of 5-week ahead COVID-19 case growth (**32.5**%) and the sample mean of daily high temperature (*≈* **79.8***°*), this implies an elasticity of **3.7**. In other words, a 10% increase in daily high temperature for a week (from **79.8***°* to **87.8***°* would predict **37**% lower case growth 5 weeks later (from **32.5**% to **20.5**%), holding mobility and the other regressors fixed.

The impact of temperature on growth in deaths becomes statistically significant one week later than for case growth, at 2 weeks ahead, and lasts longer, up to 8 weeks ahead. This result is likely explained by the fact that COVID-19 deaths have been shown to lag cases by at least one week.

Turning to precipitation and snowfall, neither is found to have a significant effect on deaths growth, but precipitation appears to have a small negative effect on COVID-19 cases 1 to 2 weeks ahead.

In sum, holding mobility fixed, temperature is found to have a negative and significant direct effect on COVID-19 cases and deaths for up to 8 weeks ahead. Precipitation may reduce growth in cases 1 to 2 weeks ahead, but it has no significant effect on case growth.

### 4.3 Dynamic Impacts of Mobility on COVID-19

I now arrive at the central results of the paper, the estimated impulse response functions for COVID-19 cases and deaths with respect to mobility shocks. These IRFs are the sequence of 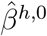 from estimating equation 2 by OLS for each weekly horizon from *h* = 1 to 10 weeks ahead. The IRFs are estimated separately for each of the alternative measures of mobility. The results for case growth are plotted in Figure 6. They reveal two general findings. First, overall mobility, as measured either by the Mobility & Engagement Index or Google Mobility’s time spent away from home, has a large positive and significant effect on case growth starting about 4 weeks ahead. The effects persists through 8 weeks ahead for the MEI and at least 10 weeks ahead for time spent away from home. The peak effect generally is around 7 weeks ahead. The effects are quantitatively large. For the MEI, the peak coefficient is **0.499**. That magnitude implies that a 1 percentage point increase (decrease) in the MEI results in a **0.5** percentage point increase (decrease) in case growth. Evaluated at sample means, this effect implies an elasticity of **0.85**, meaning that a 10% increase in mobility raises cases growth 7 weeks ahead by **8.5**%. The analogous elasticity for time spent away from home is **1.44**.

**Figure 6:**
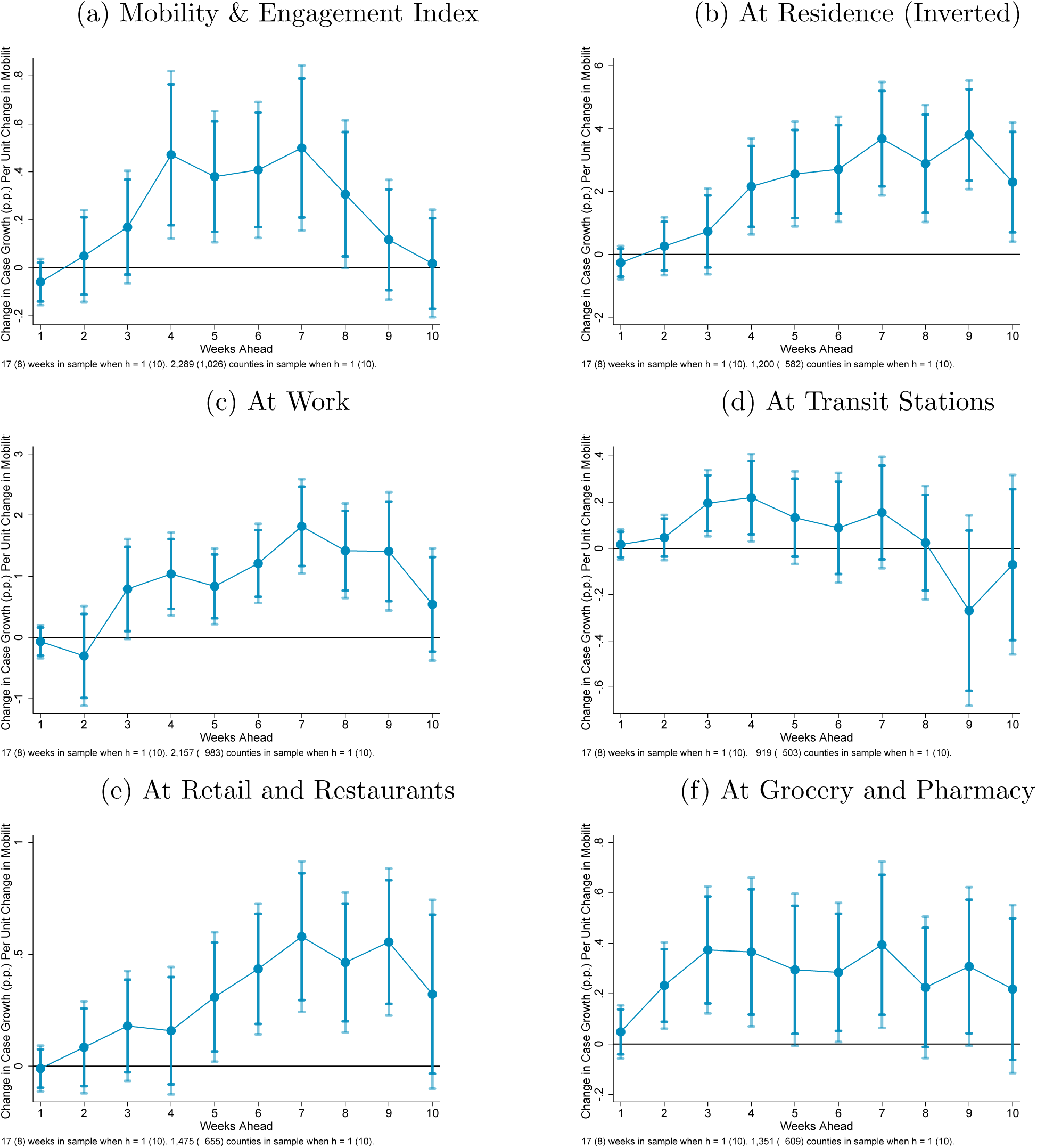
Dynamic Impacts of Mobility on COVID-19 Case Growth Impulse Response Functions Estimated by Panel Linear Projections Note: Estimates of equation 2 in the text using panel local projections regressions. Shaded regions are 90% and 95% confidence intervals.

Second, the positive effect of mobility on case growth is apparent across a variety of types of mobility. In particular, I find large and statistically significant effects for time spent at workplaces, at transit stations, at retail & restaurants, and at grocery & pharmacy.

The IRFs of mobility shocks for growth in COVID-19 deaths are shown in Figure 7. As with cases, overall mobility, measured by either the MEI or time spent away from home, has a positive and statistically significant effect on growth in deaths (see panels (a) and (b)). However, the effects do not show up until much later, around 7 weeks ahead. They appear to be continuing to increase as far as 10 weeks out. Recall that for cases the peak effect was found to be around 7 weeks ahead. As with cases, I also find significant positive effects on deaths from the specific types of mobility. For time spent at workplaces, there is actually a negative effect on deaths 1 to 3 weeks. This could potentially reflect the beneficial impact on the outcome of COVID-19 patients of healthcare and other essential workers spending more time working. At any rate, the effect turns positive 9 to 10 weeks ahead. Time spent at transit stations, at retail & restaurants, and at grocery & pharmacy also are found to increase growth in deaths at later horizons, especially 9 to 10 weeks ahead.

**Figure 7:**
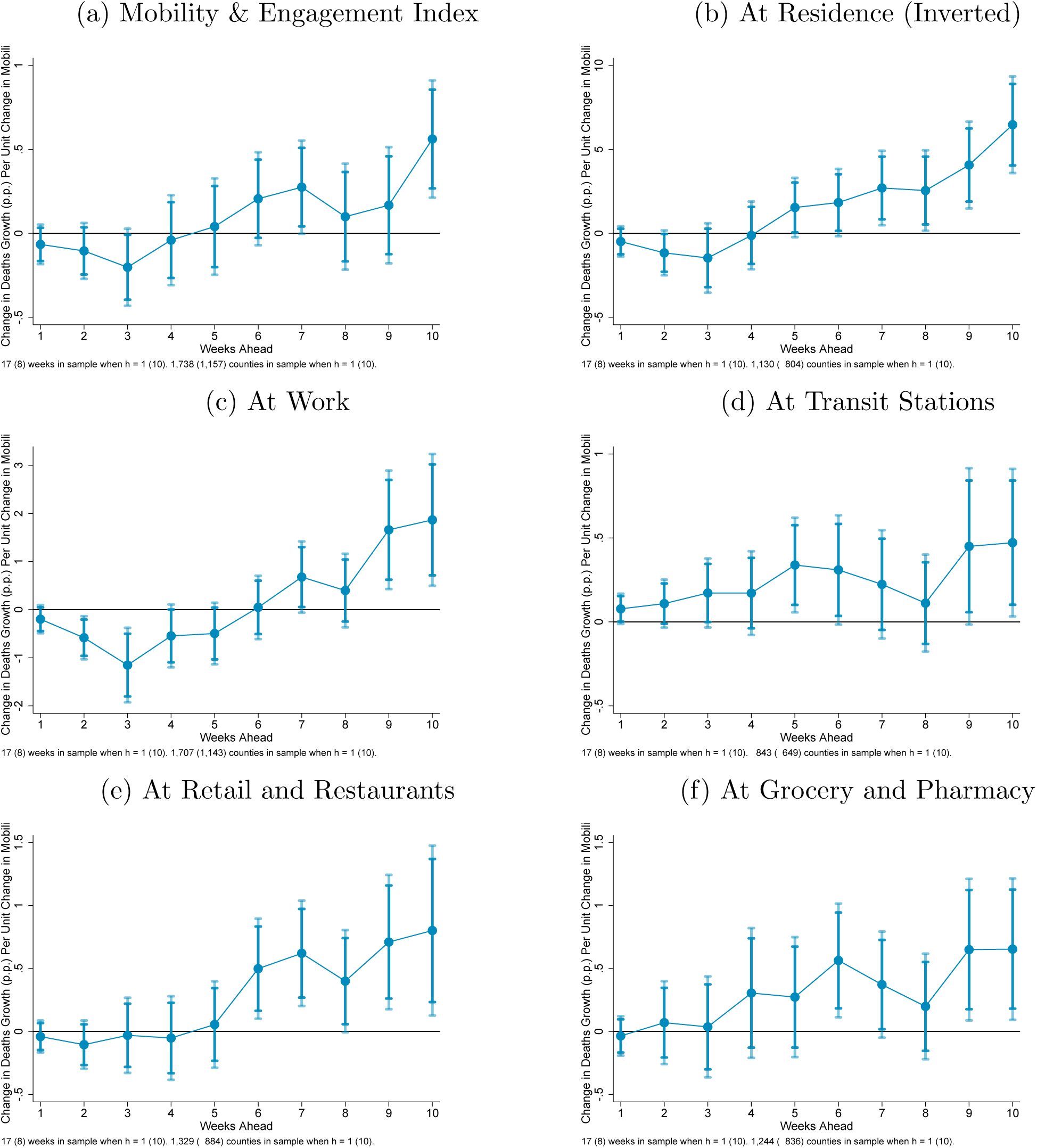
Dynamic Impacts of Mobility on COVID-19 Deaths Growth Impulse Response Functions Estimated by Panel Linear Projections Note: Estimates of equation 2 in the text using panel local projections regressions. Shaded regions are 90% and 95% confidence intervals.

The full regression results underlying these IRFs are provided, for a single selected horizon, in Tables 2 and 3. Table 2 shows the results for cases growth 7 weeks ahead and Table 3 shows the results for deaths growth 10 weeks ahead. Beyond the results for mobility already discussed, one can see in Table 2 that current case growth tends to have an insignificant effect on future case growth, though lagged case growth tends to negatively predict future case growth. Current testing growth tends to have a positive coefficient, but it is only statistically significant in 1 of the 7 specifications.

**Table 2:** Effects on 7-Week Ahead Cases Growth

|  | (1) | (2) | (3) | (4) | (5) | (6) |
| --- | --- | --- | --- | --- | --- | --- |
|  | MEI | At Home (Inv.) | At Work | At Transit | At Retail & Rest. | At Grocery & Pharmacy |
| Mobility | 0.499***<br>(0.175) | 3.674***<br>(0.919) | 1.817***<br>(0.393) | 0.155<br>(0.123) | 0.579***<br>(0.172) | 0.394**<br>(0.168) |
| L.Mobility | 0.176<br>(0.128) | 1.948**<br>(0.768) | 0.901***<br>(0.284) | -0.124<br>(0.129) | 0.163<br>(0.129) | 0.0651<br>(0.126) |
| L2.Mobility | 0.264**<br>(0.116) | 1.881**<br>(0.728) | 1.352***<br>(0.295) | 0.0849<br>(0.137) | 0.250<br>(0.155) | 0.0435<br>(0.148) |
| Cases growth rate | -0.101<br>(0.0918) | 0.0352<br>(0.0626) | -0.110<br>(0.0897) | 0.179*<br>(0.104) | 0.144**<br>(0.0682) | 0.0922<br>(0.0746) |
| L.Cases growth rate | -0.174***<br>(0.0466) | -0.167***<br>(0.0526) | -0.179***<br>(0.0428) | -0.193***<br>(0.0617) | -0.174***<br>(0.0546) | -0.177***<br>(0.0650) |
| L2.Cases growth rate | -0.240***<br>(0.0514) | -0.216***<br>(0.0506) | -0.191***<br>(0.0398) | -0.285***<br>(0.0622) | -0.269***<br>(0.0569) | -0.271***<br>(0.0542) |
| Testing growth rate | 133.0*<br>(80.66) | 59.61<br>(72.71) | 66.55<br>(81.30) | 129.4<br>(91.35) | 85.12<br>(71.92) | 66.41<br>(75.44) |
| Max daily temp | -0.360**<br>(0.170) | -1.039***<br>(0.192) | -0.672***<br>(0.165) | -0.677***<br>(0.215) | -0.772***<br>(0.189) | -0.763***<br>(0.200) |
| Precipitation | 0.180<br>(0.178) | 0.160<br>(0.207) | 0.0234<br>(0.149) | 0.173<br>(0.244) | 0.0801<br>(0.199) | 0.0367<br>(0.210) |
| Snowfall | -1.486<br>(2.055) | 0.623<br>(1.620) | -1.095<br>(1.729) | -3.239<br>(2.363) | -2.092<br>(1.568) | -1.561<br>(1.719) |
| Observations | 8604 | 4838 | 8243 | 4136 | 5365 | 5005 |
| Adjusted $R^2$ | 0.032 | 0.058 | 0.061 | 0.023 | 0.047 | 0.027 |
| Dep. Variable Mean | 35.764 | 35.421 | 35.999 | 36.841 | 35.130 | 36.116 |
| Mobility Mean | -61.148 | -13.838 | -33.087 | -24.027 | -23.017 | -2.875 |
| Elasticity at means | .85 | 1.44 | 1.67 | .1 | .38 | .03 |

\* $p < 0.10$ , \*\* $p < 0.05$ , \*\*\* $p < 0.01$

**Table 3:** Effects on 10-Week Ahead Deaths Growth

|  | (1) | (2) | (3) | (4) | (5) | (6) |
| --- | --- | --- | --- | --- | --- | --- |
|  | MEI | At Home (Inv.) | At Work | At Transit | At Retail & Rest. | At Grocery & Pharmacy |
| Mobility | 0.562***<br>(0.178) | 6.469***<br>(1.468) | 1.866***<br>(0.699) | 0.472**<br>(0.224) | 0.801**<br>(0.344) | 0.654**<br>(0.287) |
| L.Mobility | 0.110<br>(0.169) | 2.912**<br>(1.328) | 0.456<br>(0.645) | -0.0574<br>(0.230) | 0.437*<br>(0.243) | 0.188<br>(0.278) |
| L2.Mobility | 0.402*<br>(0.236) | 1.122<br>(1.378) | 0.549<br>(0.534) | 0.120<br>(0.231) | -0.230<br>(0.255) | 0.178<br>(0.214) |
| Deaths growth rate | -0.208***<br>(0.0545) | -0.284***<br>(0.0725) | -0.200***<br>(0.0555) | -0.301***<br>(0.0715) | -0.286***<br>(0.0698) | -0.304***<br>(0.0707) |
| L.Deaths growth rate | -0.111***<br>(0.0405) | -0.179***<br>(0.0532) | -0.106**<br>(0.0418) | -0.234***<br>(0.0581) | -0.146***<br>(0.0528) | -0.174***<br>(0.0538) |
| L2.Deaths growth rate | -0.0930***<br>(0.0265) | -0.116***<br>(0.0361) | -0.0988***<br>(0.0276) | -0.107***<br>(0.0393) | -0.0947***<br>(0.0334) | -0.106***<br>(0.0352) |
| Cases growth rate | 0.194*<br>(0.100) | 0.200*<br>(0.117) | 0.189*<br>(0.106) | 0.223*<br>(0.118) | 0.168<br>(0.114) | 0.102<br>(0.119) |
| Testing growth rate | 5.876<br>(99.52) | -3.119<br>(123.8) | -26.37<br>(101.9) | 96.76<br>(138.3) | 30.66<br>(122.2) | 5.657<br>(132.1) |
| Max daily temp | 0.219<br>(0.203) | 0.0180<br>(0.230) | 0.196<br>(0.208) | 0.289<br>(0.251) | 0.296<br>(0.252) | 0.168<br>(0.239) |
| Precipitation | 0.205<br>(0.272) | 0.0401<br>(0.305) | 0.185<br>(0.272) | 0.117<br>(0.385) | -0.0228<br>(0.293) | -0.112<br>(0.298) |
| Snowfall | -1.806<br>(1.323) | 0.376<br>(1.666) | -1.686<br>(1.364) | -0.888<br>(1.623) | -0.666<br>(1.525) | -1.259<br>(1.478) |
| Observations | 4577 | 3337 | 4516 | 2732 | 3613 | 3438 |
| Adjusted $R^2$ | 0.023 | 0.044 | 0.020 | 0.032 | 0.027 | 0.026 |
| Dep. Variable Mean | 23.040 | 25.858 | 23.167 | 27.427 | 25.198 | 25.685 |
| Mobility Mean | -75.026 | -15.838 | -38.009 | -30.933 | -30.365 | -6.607 |
| Elasticity at means | 1.83 | 3.96 | 3.06 | .53 | .97 | .17 |

\* $p < 0.10$ , \*\* $p < 0.05$ , \*\*\* $p < 0.01$

The results are qualitatively similar for future deaths growth (Table 3), though both current and lagged deaths growth are predictive of future deaths growth, with negative signs. This is consistent with mean reversion in death growth rates.

As noted in Section 3.2, one can also estimate the dynamic effects of mobility on *cumulative* growth in cases or deaths out to any given horizon. Table 4 report the effects of current mobility and other variables on cumulative growth in cases over the subsequent 8 weeks. 90% confidence intervals are shown below each coefficient. The 8-week ahead cumulative effect is positive in all regressions. For cumulative growth in cases, it is statistically significant (below the 10% level) for all measures of mobility except time spent at transit stations. The effects of mobility are, in general, quantitatively large. In particular, the coefficient on time spent away from home in Table 4 of **27.35** implies that a one percentage point increase (decrease) in that mobility measure leads to an increase (decrease) in cumulative case growth over the subsequent 8 weeks of about **27** percentage points, or about 12% of average 8-week case growth in the sample (**225.95**%, shown at the bottom of the table). The effect magnitudes, expressed as elasticities, are shown at the bottom of the table. The **27.35** coefficient on time spent away from home implies an elasticity of **1.78**, implying that a 10% increase in mobility leads to **17.8**% higher cumulative case growth 8 weeks ahead.

**Table 4:** Estimates of Cumulative Effect on Cases Growth over Subsequent 8 Weeks

|  | (1) | (2) | (3) | (4) | (5) | (6) |
| --- | --- | --- | --- | --- | --- | --- |
|  | MEI | At Home (Inv.) | At Work | At Transit | At Retail & Rest. | At Grocery & Pharmacy |
| Mobility | 2.323**<br>[0.597,4.048] | 27.35***<br>[17.96,36.73] | 15.52***<br>[10.33,20.72] | 0.797<br>[-0.392,1.985] | 3.650***<br>[1.782,5.518] | 2.953***<br>[1.277,4.629] |
| L.Mobility | 0.736<br>[-1.110,2.583] | 31.01***<br>[21.22,40.79] | 12.57***<br>[7.016,18.12] | 0.156<br>[-1.095,1.408] | 3.756***<br>[2.071,5.442] | 1.542<br>[-0.121,3.205] |
| L2.Mobility | 3.237***<br>[2.026,4.449] | 23.48***<br>[14.13,32.83] | 15.39***<br>[10.95,19.82] | 1.592*<br>[0.0711,3.113] | 0.339<br>[-1.455,2.133] | 0.436<br>[-1.512,2.384] |
| Cases growth rate | 0.110<br>[-0.981,1.201] | 1.638***<br>[0.818,2.458] | 0.117<br>[-0.806,1.040] | 2.254***<br>[1.068,3.440] | 2.707***<br>[1.713,3.702] | 2.266***<br>[1.291,3.241] |
| L.Cases growth rate | -2.473***<br>[-3.201,-1.745] | -2.154***<br>[-2.840,-1.468] | -2.649***<br>[-3.288,-2.011] | -1.815***<br>[-2.414,-1.216] | -1.998***<br>[-2.731,-1.265] | -2.217***<br>[-3.102,-1.333] |
| L2.Cases growth rate | -2.593***<br>[-3.239,-1.948] | -2.612***<br>[-3.137,-2.086] | -2.245***<br>[-2.725,-1.764] | -2.933***<br>[-3.581,-2.286] | -3.196***<br>[-3.802,-2.591] | -3.351***<br>[-3.942,-2.759] |
| Testing growth rate | 1647.6***<br>[663.6,2631.7] | 1748.8**<br>[610.4,2887.3] | 1199.6**<br>[253.9,2145.2] | 2264.9***<br>[995.7,3534.0] | 1993.4***<br>[869.2,3117.6] | 2024.4***<br>[780.0,3268.8] |
| Max daily temp | -0.203<br>[-1.983,1.578] | -5.586***<br>[-7.359,-3.812] | -2.212**<br>[-3.786,-0.638] | -2.148**<br>[-3.941,-0.354] | -3.494***<br>[-5.334,-1.653] | -3.654***<br>[-5.481,-1.827] |
| Precipitation | 1.401<br>[-0.465,3.267] | 0.279<br>[-1.338,1.897] | 1.521<br>[-0.299,3.342] | 0.152<br>[-2.025,2.329] | -0.0930<br>[-1.722,1.536] | -0.437<br>[-2.238,1.365] |
| Snowfall | -25.72*<br>[-49.91,-1.530] | 11.23<br>[-8.438,30.90] | -20.68<br>[-42.60,1.234] | -7.549<br>[-29.14,14.04] | -12.56<br>[-31.55,6.430] | -3.282<br>[-21.60,15.04] |
| Observations | 6945 | 3933 | 6664 | 3414 | 4374 | 4073 |
| Adjusted $R^2$ | 0.074 | 0.233 | 0.149 | 0.109 | 0.169 | 0.146 |
| Dep. Variable Mean | 238.962 | 225.945 | 237.935 | 236.222 | 224.847 | 227.737 |
| Mobility Mean | -65.696 | -14.719 | -35.043 | -26.805 | -25.710 | -4.272 |
| Elasticity at means | .64 | 1.78 | 2.29 | .09 | .42 | .06 |

\* $p < 0.10$ , \*\* $p < 0.05$ , \*\*\* $p < 0.01$

The results for cumulative growth in COVID-19 deaths 10 weeks ahead are provided in Table 5. All measures of mobility are found to have a positive and significant impact on deaths. The elasticities, shown at the bottom of the table, are higher than the corresponding elasticities for cumulative case growth in Table 4. For instance, the elasticity with respect to time spent away from home is found to be **3.25**.

**Table 5:** Estimates of Cumulative Effect on Deaths Growth over Subsequent 10 Weeks

|  | (1) | (2) | (3) | (4) | (5) | (6) |
| --- | --- | --- | --- | --- | --- | --- |
|  | MEI | At Home (Inv.) | At Work | At Transit | At Retail & Rest. | At Grocery & Pharmacy |
| Mobility | 6.770***<br>[2.963,10.58] | 69.03***<br>[38.57,99.48] | 22.85***<br>[10.51,35.19] | 7.642**<br>[2.251,13.03] | 16.11***<br>[8.616,23.60] | 13.53***<br>[6.654,20.41] |
| L.Mobility | 3.638*<br>[0.348,6.929] | 56.91***<br>[24.04,89.78] | 22.00***<br>[9.488,34.51] | -1.101<br>[-5.563,3.362] | 7.569***<br>[3.066,12.07] | 3.453<br>[-2.015,8.922] |
| L2.Mobility | 12.39***<br>[5.645,19.13] | 85.19***<br>[38.62,131.8] | 29.34***<br>[13.25,45.43] | 2.362<br>[-2.666,7.391] | 2.382<br>[-3.108,7.873] | 5.344*<br>[0.667,10.02] |
| Deaths growth rate | -3.732***<br>[-5.085,-2.378] | -4.768***<br>[-6.513,-3.024] | -3.597***<br>[-4.947,-2.248] | -5.329***<br>[-7.492,-3.166] | -4.907***<br>[-6.633,-3.181] | -4.791***<br>[-6.561,-3.021] |
| L.Deaths growth rate | -2.995***<br>[-3.701,-2.289] | -3.993***<br>[-4.975,-3.010] | -2.901***<br>[-3.601,-2.201] | -4.835***<br>[-6.143,-3.528] | -3.786***<br>[-4.737,-2.835] | -4.040***<br>[-5.073,-3.007] |
| L2.Deaths growth rate | -2.151***<br>[-2.605,-1.698] | -2.584***<br>[-3.213,-1.954] | -2.204***<br>[-2.684,-1.723] | -2.996***<br>[-3.711,-2.280] | -2.173***<br>[-2.738,-1.607] | -2.501***<br>[-3.122,-1.880] |
| Cases growth rate | 3.551***<br>[2.010,5.092] | 3.622***<br>[1.640,5.604] | 3.456***<br>[1.903,5.008] | 3.800**<br>[1.018,6.582] | 4.356***<br>[2.382,6.330] | 3.161**<br>[1.104,5.217] |
| Testing growth rate | 2483.0<br>[-164.6,5130.5] | 2717.3<br>[-563.1,5997.7] | 1720.0<br>[-967.6,4407.6] | 3601.1<br>[-393.5,7595.7] | 3137.3<br>[-150.4,6425.1] | 3050.5<br>[-503.2,6604.2] |
| Max daily temp | 5.329*<br>[0.442,10.22] | 2.873<br>[-2.528,8.273] | 4.469<br>[-0.216,9.153] | 9.323***<br>[3.472,15.17] | 7.035*<br>[0.357,13.71] | 4.432<br>[-1.300,10.16] |
| Precipitation | 0.114<br>[-4.703,4.932] | -2.002<br>[-7.514,3.511] | -0.420<br>[-5.265,4.425] | 0.479<br>[-5.758,6.716] | -1.990<br>[-7.401,3.422] | -3.532<br>[-8.840,1.776] |
| Snowfall | -5.460<br>[-39.69,28.77] | 17.02<br>[-20.36,54.41] | -0.928<br>[-32.67,30.82] | 22.77<br>[-12.83,58.36] | 21.61<br>[-11.61,54.82] | 7.075<br>[-27.76,41.91] |
| Observations | 4577 | 3337 | 4516 | 2732 | 3613 | 3438 |
| Adjusted $R^2$ | 0.134 | 0.174 | 0.136 | 0.128 | 0.149 | 0.125 |
| Dep. Variable Mean | 290.332 | 336.334 | 292.321 | 351.414 | 323.753 | 332.122 |
| Mobility Mean | -75.026 | -15.838 | -38.009 | -30.933 | -30.365 | -6.607 |
| Elasticity at means | 1.75 | 3.25 | 2.97 | .67 | 1.51 | .27 |

\* $p < 0.10$ , \*\* $p < 0.05$ , \*\*\* $p < 0.01$

Lastly, it is interesting to consider the adjusted-*R*^2^’s, shown at the bottom of Tables 4 and 5. The regressors in the model, including the county and week fixed effects, explain as much as **23**% of the variation in cumulative case growth 8 weeks ahead and as much as **17**% of variation in cumulative deaths growth 10 weeks ahead. In other words, **77** to **83**% of the variation in these COVID-19 outcomes is not readily explainable by time-invariant county characteristics, common national time-varying factors, recent case and testing growth, weather, and mobility.

In sum, overall mobility is found to have a large positive and long-lasting effect on subsequent growth in COVID-19 cases and deaths. The effects become apparent around 3-4 weeks ahead for cases and around 5-6 weeks ahead for deaths. The peak effect occurs around 7 weeks ahead for cases and 10 or more weeks ahead for deaths. Looking across subcategories of mobility, the effects are clearest for time spent at workplaces and retail & recreation, though there is also evidence of adverse effects from time spent at transit stations and at grocery & pharmacy locations.

## 5 Extensions

### 5.1 Heterogeneous Effects of Mobility

The results presented thus far have focused on the average treatment effect of mobility on COVID-19 outcomes. The true impact of mobility is likely to vary across time and space, depending on certain local conditions at the time and demographic and other fixed characteristics of the locality. One particular condition that could be expected to amplify the impact of mobility on COVID-19 spread is the local transmission rate of the virus, which is commonly denoted *R*_*t*_ in epidemiological SIR models. In these models, new infections are the result of an interaction between the contact rate – the rate at which individuals come into contact with other, potentially infectious, individuals – and the transmission rate, which is the average number of individuals who contract the virus from exposure to one infected person. Mobility can be thought of as a proxy for the unobserved contact rate at a particular time and place. To proxy for the transmission rate, I follow the approach of Xu, Rahmandad, Gupta, DiGennaro, Ghaffarzadegan, Amini, and Jalali (2020). They approximate *R*_*t*_ by the daily case count divided by its average over the prior 20 days. The latter is a proxy for the number of potentially infectious individuals per day, assuming an average interval from exposure to recovery (or death) of 20 days.

To assess the dependence of mobility’s impacts on COVID-19 cases and deaths to the local transmission rate, I split the sample into county-week observations for which *R*_*t*_ <= 1 and for which *R*_*t*_ > 1. 1 is a critical threshold for the transmission rate, with virus spread expected to asymptote to zero when *R*_*t*_ is below one and expected to increase indefinitely when *R*_*t*_ is above one. I then estimate the mobility IRFs for subsequent growth in cases and deaths separately for each subsample.

The results are shown in Figure 8. Panels (a) and (b) show the mobility IRFs for growth in cases, while panels (c) and (d) show the IRFs for growth in deaths. Mobility here is measured using time spent away from home. IRFs using the other mobility measures yield qualitatively similar results and are provided in Appendix Figures A1–A3. The results show quite clearly that the deleterious effects of mobility on COVID-19 outcomes are far greater when the local transmission rate is above one. When *R*_*t*_ is below one, mobility increases (decreases) have a small positive (negative) effect on subsequent cases but virtually no effect on deaths. When *R*_*t*_ is above one, mobility changes have a much larger effect on subsequent cases and have a large, long-lasting impact on future deaths. The results in Appendix Figures A1–A3 show that these patterns are robust across all measures of mobility.

**Figure 8:**
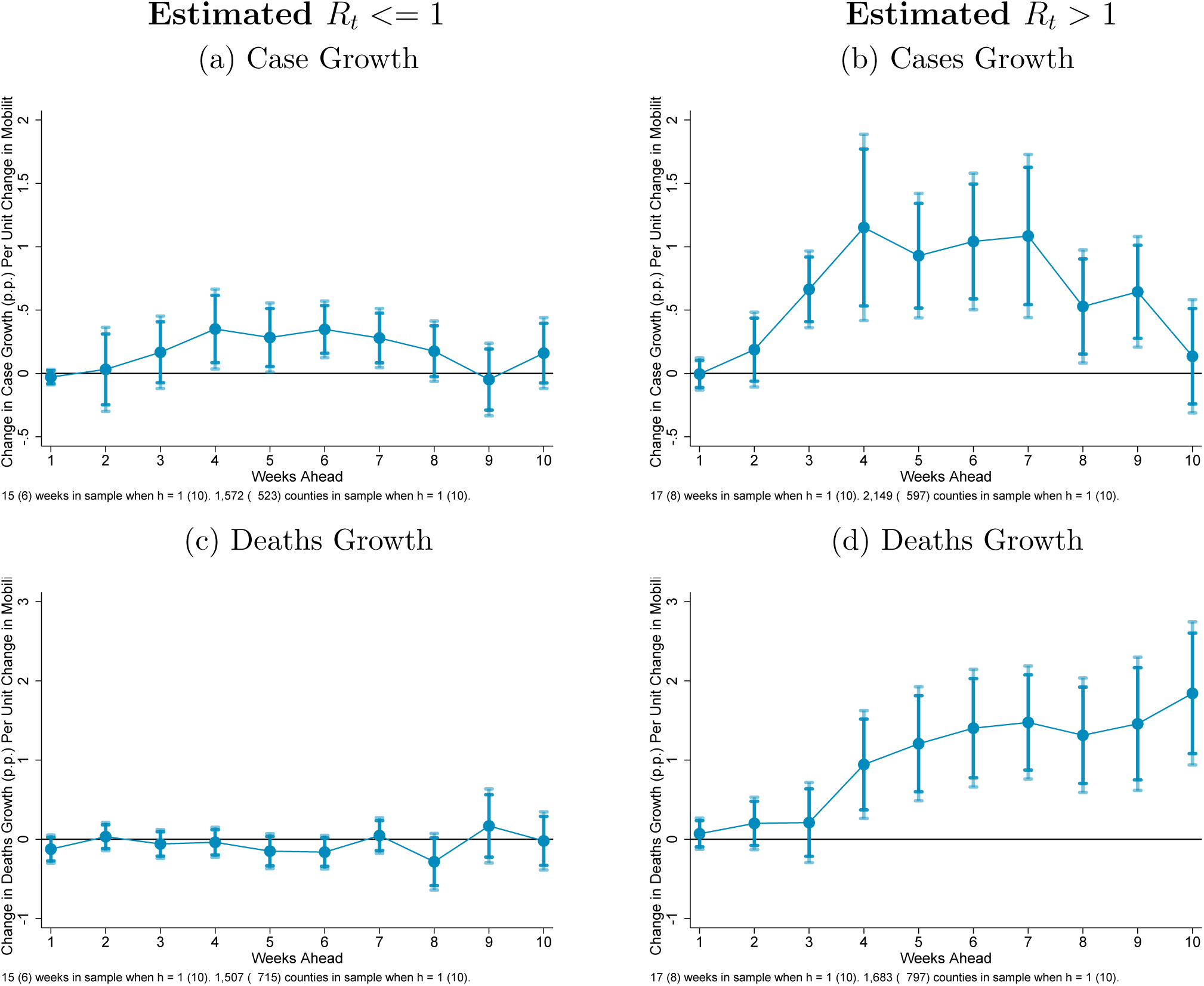
Dynamic Impacts of Mobility on Growth in COVID-19 Outcomes Note: Estimates of equation 2 in the text using panel local projections regressions. Mobility measured by the Mobility & Engagement Index. Shaded regions are 90% and 95% confidence intervals.

I next consider whether mobility’s effects vary across different types of counties – that is, whether they are heterogeneous with respect to certain fixed county characteristics. Specifically, I consider heterogeneity along the following dimensions: the population share in nursing homes, the share 60 years old and over, the share that identify as black, the share that identify as hispanic, and average household size. For each dimension I construct an indicator that is 1 if the variable is above the median value over all counties and 0 otherwise. I then interact the indicator with current and lagged mobility, and add the interactions to the local projections specification (equation 2). I plot the implied separate IRFs for high versus low. The results, using mobility measured by time spent away from home, are shown in Figure 9 for cases growth and in Figure 10 for deaths growth. The estimated IRFs are found not to differ much across these dimensions for cases growth. However, there are some important differences for deaths growth. First, mobility increases appear to be more deleterious for COVID-19 fatality in counties with a higher share of the population over 60. This likely reflects that well-known fact that COVID-19 infections are more likely to be fatal for older individuals. Thus, increases in mobility in counties with older populations likely lead to increased coronavirus exposure by older individuals. One does not see a differential effect of mobility for counties with relatively high populations shares in nursing homes. This may be because mobility increases in higher nursing home share counties primarily reflect the mobility behavior of the vast majority of the population *not* in nursing homes given that even above-median counties still have a fairly low nursing home share. (The median is only 2%.)

**Figure 9:**
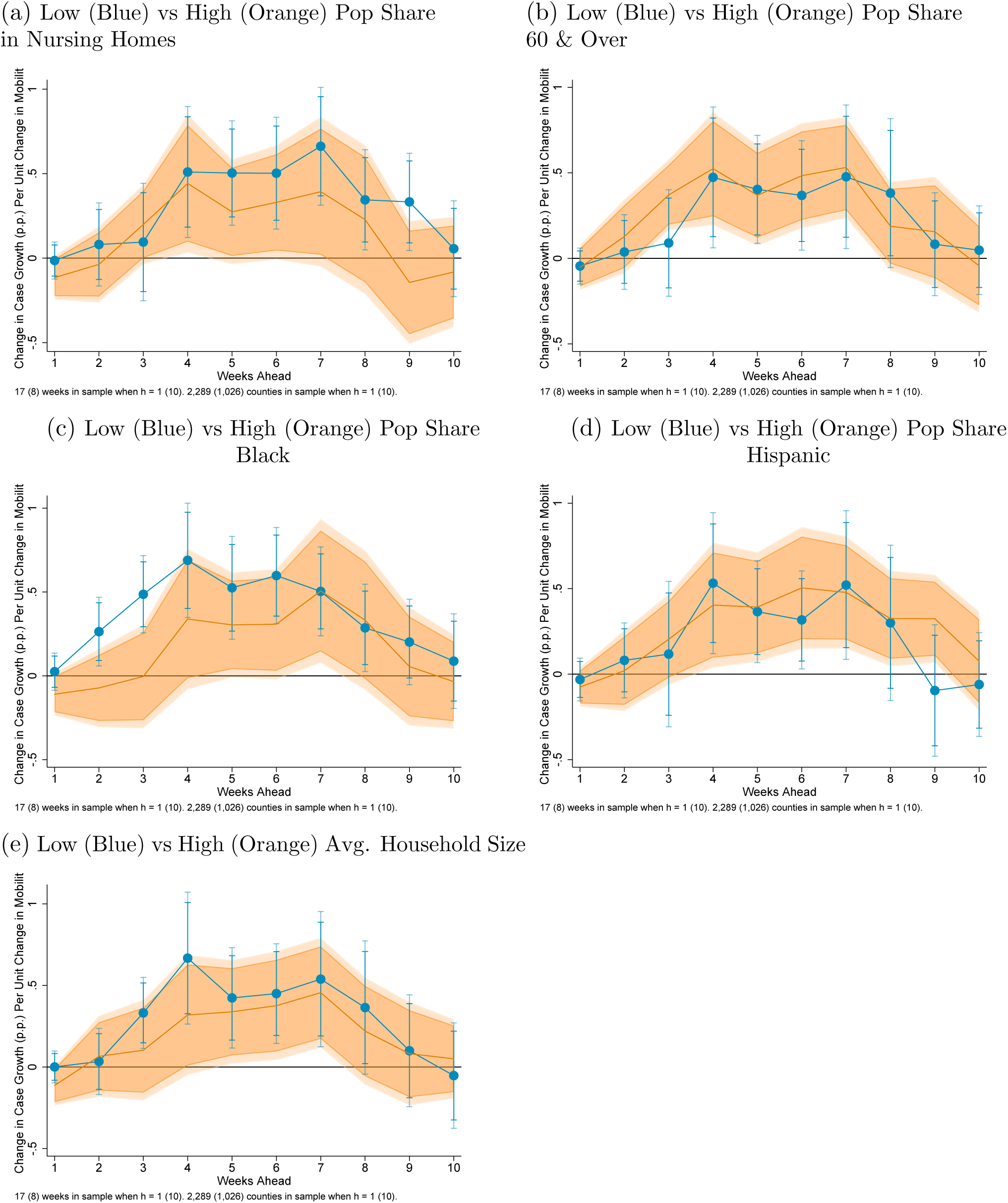
Dynamic Impacts of Mobility on COVID-19 Cases Growth Note: Estimates of equation 2 in the text using panel local projections regressions. Shaded regions are 90% and 95% confidence intervals.

**Figure 10:**
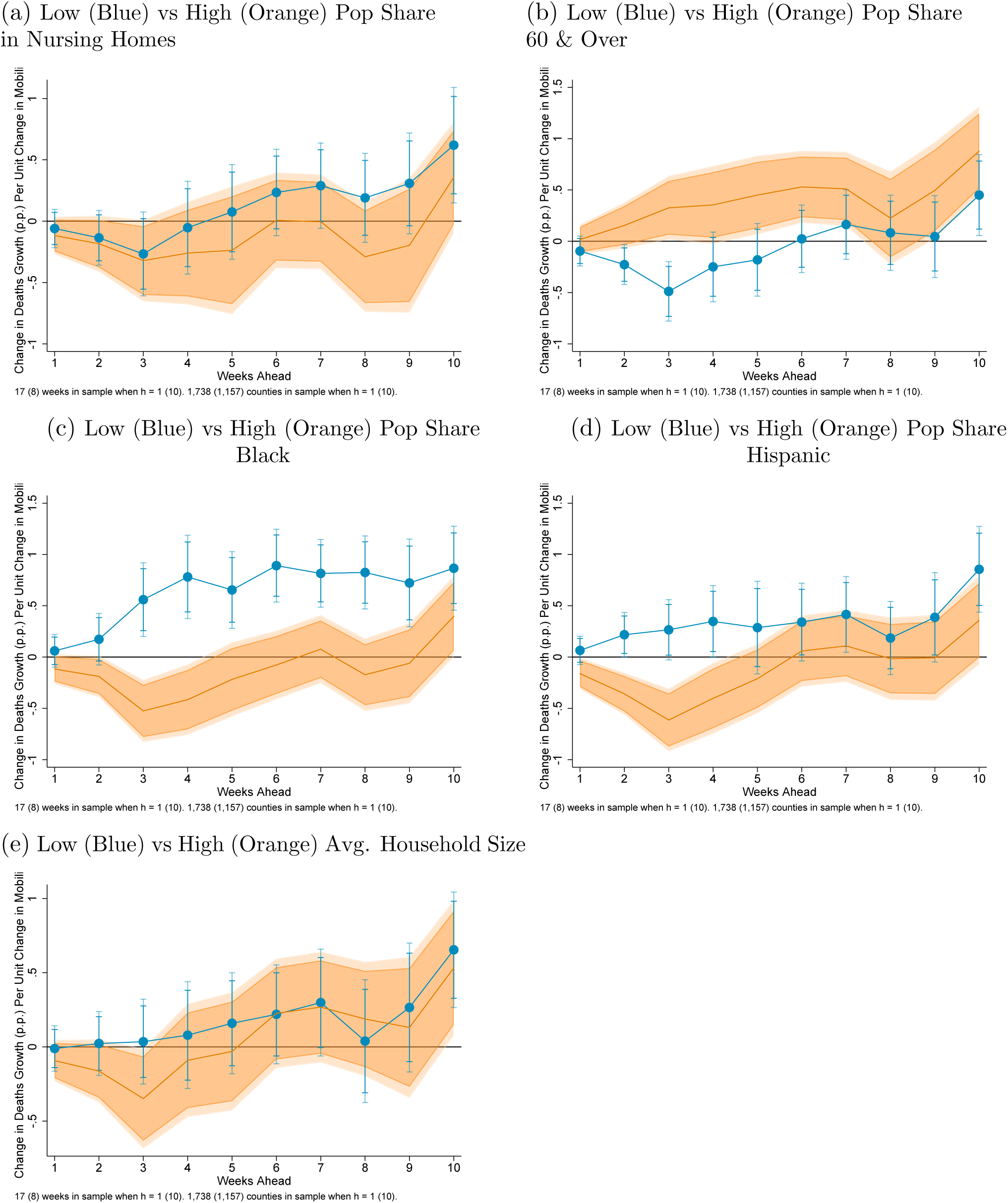
Dynamic Impacts of Mobility on COVID-19 Deaths Growth - Weekly Note: Estimates of equation 2 in the text using panel local projections regressions. Shaded regions are 90% and 95% confidence intervals.

Surprisingly, mobility appears to have a lower impact of subsequent deaths in counties with higher shares of black or hispanic populations. That is, despite the fact, documented in prior research, that counties with higher black and hispanic population shares tend to have higher COVID-19 cases and deaths, the marginal impact of mobility on deaths appears to go the other way. One possible explanation is that when places reduce mobility sharply – e.g., in accordance with shelter-in-place orders – demand for essential services such as food and parcel delivery rises. The workforce for such essential services has been shown to be disproportionately comprised of black and hispanic individuals. Thus, it is possible that overall decreases (increases) in mobility may actually lead to increases (decreases) in virus exposure for these groups.

### 5.2 The Role of Public Health Policies

To the extent that public health policies, generally known as non-pharmaceutical interventions (NPIs), affect COVID-19 outcomes, their effects are likely to work primarily through the channel of affecting individuals’ mobility/social-distancing behavior. Indeed, I find that the number of NPIs in place in a county (or state of the county where county NPI data is unavailable) has a strong reducing effect on mobility when estimating equation 1 with the number of NPIs added as a regressor. This is shown in Table 6, which provides the results from daily panel fixed effects regressions of each measure of mobility on the number of local NPIs in place as well the set of weather variables used in Table 1. I find that NPIs have a positive and and statistically significant effect on all measures of mobility. Thus, NPIs should indirectly reduce COVID-19 cases and deaths through the channel of reducing mobility, which in turn reduces cases and deaths.

**Table 6:** Effect of Non-Pharmaceutical Interventions on Mobility

|  | (1) | (2) | (3) | (4) | (5) | (6) |
| --- | --- | --- | --- | --- | --- | --- |
|  | MEI | At Home (Inverted) | At Work | At Transit | At Retail & Rest. | At Grocery & Pharmacy |
| Number of NPIs in place (out of 10) | -3.069***<br>(0.257) | -0.716***<br>(0.0524) | -1.244***<br>(0.0991) | -2.580***<br>(0.347) | -2.111***<br>(0.209) | -1.446***<br>(0.190) |
| Max Daily Temp – Weekday | 0.0621***<br>(0.0139) | 0.0652***<br>(0.00306) | 0.0555***<br>(0.00580) | 0.191***<br>(0.0182) | 0.193***<br>(0.0130) | 0.181***<br>(0.0118) |
| Max Daily Temp – Weekend | -0.0375**<br>(0.0186) | -0.00551<br>(0.00518) | -0.0400***<br>(0.00882) | 0.0147<br>(0.0237) | 0.112***<br>(0.0168) | 0.141***<br>(0.0154) |
| Precipitation – Weekday | -0.137***<br>(0.0121) | -0.0215***<br>(0.00293) | -0.0115***<br>(0.00414) | -0.0390***<br>(0.00915) | -0.0342***<br>(0.00962) | -0.0312***<br>(0.00883) |
| Precipitation – Weekend | -0.155***<br>(0.0213) | -0.0331***<br>(0.00704) | -0.0232**<br>(0.0107) | -0.0907***<br>(0.0224) | -0.0651***<br>(0.0194) | -0.0775***<br>(0.0202) |
| Snowfall – Weekday | -0.723***<br>(0.176) | -0.0845***<br>(0.0317) | -0.180***<br>(0.0535) | -0.222*<br>(0.116) | -0.351***<br>(0.0882) | -0.393***<br>(0.0848) |
| Snowfall – Weekend | -0.474**<br>(0.194) | -0.273***<br>(0.0530) | -0.278***<br>(0.0908) | -0.988***<br>(0.271) | -0.529***<br>(0.149) | -0.404***<br>(0.148) |
| Observations | 417096 | 151365 | 316480 | 127135 | 226524 | 211709 |
| Adjusted $R^2$ | 0.864 | 0.915 | 0.894 | 0.755 | 0.811 | 0.669 |
| Earliest date | 22jan2020 | 15feb2020 | 15feb2020 | 15feb2020 | 15feb2020 | 15feb2020 |
| Latest date | 11jul2020 | 15jul2020 | 15jul2020 | 15jul2020 | 15jul2020 | 15jul2020 |
| # of days | 171 | 152 | 152 | 152 | 152 | 152 |
| # of counties | 2679 | 1408 | 2486 | 1046 | 2297 | 2207 |

\* $p < 0.10$ , \*\* $p < 0.05$ , \*\*\* $p < 0.01$

It is possible that NPIs have additional effects on COVID-19 outcomes through other channels such as encouraging people to wash their hands, wear masks, and stay physically distant from others even when spending time in public places. Also, empirically, NPIs could be found to have direct effects on COVID-19 outcomes if mobility is incompletely measured. I assess the effects of NPIs, holding measured mobility fixed, by adding the number of local NPIs in place as of date *t* to the estimations of equation 2 using growth in cases or deaths as the dependent variable. The results for 7-week ahead case growth are shown in Table 7. Consistent with the hypothesis that NPIs have additional effects beyond the (measured) mobility channel, I find that NPIs have a significant effect on future COVID-19 case growth nearly all specifications. The one exception is the case where mobility is measured by the MEI. The results for 10-week ahead growth in deaths are shown in 8. In contrast to the results for cases, the estimated effect of NPIs on future growth in deaths is negative but statistically insignificant in all but one specification. Thus, whether NPIs have a direct effect, outside of the mobility channel, on deaths is much less clear. In the regressions for cumulative deaths growth (Table 8), mobility has a significant effect in all specifications while NPIs are insignificant in all specifications. It is also worth noting that, comparing the results in Tables 7 and 8 to the analogous results in Tables 2 and 3 which omit NPIs, the coefficients on mobility are virtually unchanged by including NPIs.

**Table 7:** Estimates of Effect on 7-Week Ahead Cases Growth Controlling for Non-Pharmaceutical Interventions

Controlling for Non-Pharmaceutical Interventions
|  | (1) | (2) | (3) | (4) | (5) | (6) |
| --- | --- | --- | --- | --- | --- | --- |
|  | MEI | At Home (Inv.) | At Work | At Transit | At Retail & Rest. | At Grocery & Pharmacy |
| Mobility | 0.576***<br>(0.160) | 2.859***<br>(0.859) | 1.395***<br>(0.351) | 0.143<br>(0.112) | 0.489***<br>(0.174) | 0.279*<br>(0.152) |
| L.Mobility | 0.196<br>(0.130) | 2.134**<br>(0.828) | 1.410***<br>(0.280) | 0.0922<br>(0.0984) | 0.334**<br>(0.151) | 0.164<br>(0.142) |
| Cases growth rate | -0.0571<br>(0.0629) | 0.0678<br>(0.0780) | -0.0579<br>(0.0569) | 0.0448<br>(0.0905) | 0.141*<br>(0.0790) | 0.106<br>(0.0885) |
| L.Cases growth rate | -0.175***<br>(0.0582) | -0.183***<br>(0.0697) | -0.192***<br>(0.0499) | -0.0866<br>(0.0869) | -0.146**<br>(0.0699) | -0.173**<br>(0.0774) |
| Testing growth rate | 236.4***<br>(79.96) | 165.3*<br>(84.33) | 161.0**<br>(78.59) | 258.9***<br>(98.62) | 213.0**<br>(83.55) | 186.6**<br>(85.46) |
| L2.Cases growth rate | -0.284***<br>(0.0461) | -0.325***<br>(0.0572) | -0.222***<br>(0.0426) | -0.406***<br>(0.0689) | -0.375***<br>(0.0587) | -0.370***<br>(0.0621) |
| L2.Mobility | 0.334**<br>(0.129) | 2.481***<br>(0.729) | 1.018***<br>(0.267) | -0.0991<br>(0.138) | 0.0241<br>(0.145) | -0.00237<br>(0.148) |
| Max daily temp | -0.735***<br>(0.177) | -1.225***<br>(0.229) | -0.874***<br>(0.172) | -0.768***<br>(0.224) | -0.921***<br>(0.213) | -0.875***<br>(0.227) |
| Precipitation | 0.376**<br>(0.175) | 0.356<br>(0.218) | 0.317**<br>(0.157) | 0.422*<br>(0.226) | 0.343*<br>(0.205) | 0.255<br>(0.214) |
| Snowfall | -3.162<br>(2.057) | -0.715<br>(1.736) | -2.818*<br>(1.700) | -2.088<br>(1.911) | -2.551<br>(1.587) | -1.898<br>(1.680) |
| # of NPIs in place | -2.545<br>(1.555) | -4.082**<br>(1.714) | -3.056**<br>(1.481) | -4.424**<br>(1.978) | -3.990**<br>(1.691) | -5.237***<br>(1.771) |
| Observations | 9108 | 5489 | 8933 | 4575 | 6240 | 5863 |
| Adjusted $R^2$ | 0.051 | 0.076 | 0.076 | 0.042 | 0.059 | 0.044 |
| Dep. variable mean | 33.286 | 33.218 | 33.159 | 34.379 | 33.162 | 34.139 |
| Mobility mean | -58.400 | -13.159 | -31.930 | -22.520 | -20.505 | -1.238 |
| Elasticity at means | 1.01 | 1.13 | 1.34 | .09 | .3 | .01 |

\* $p < 0.10$ , \*\* $p < 0.05$ , \*\*\* $p < 0.01$

**Table 8:**
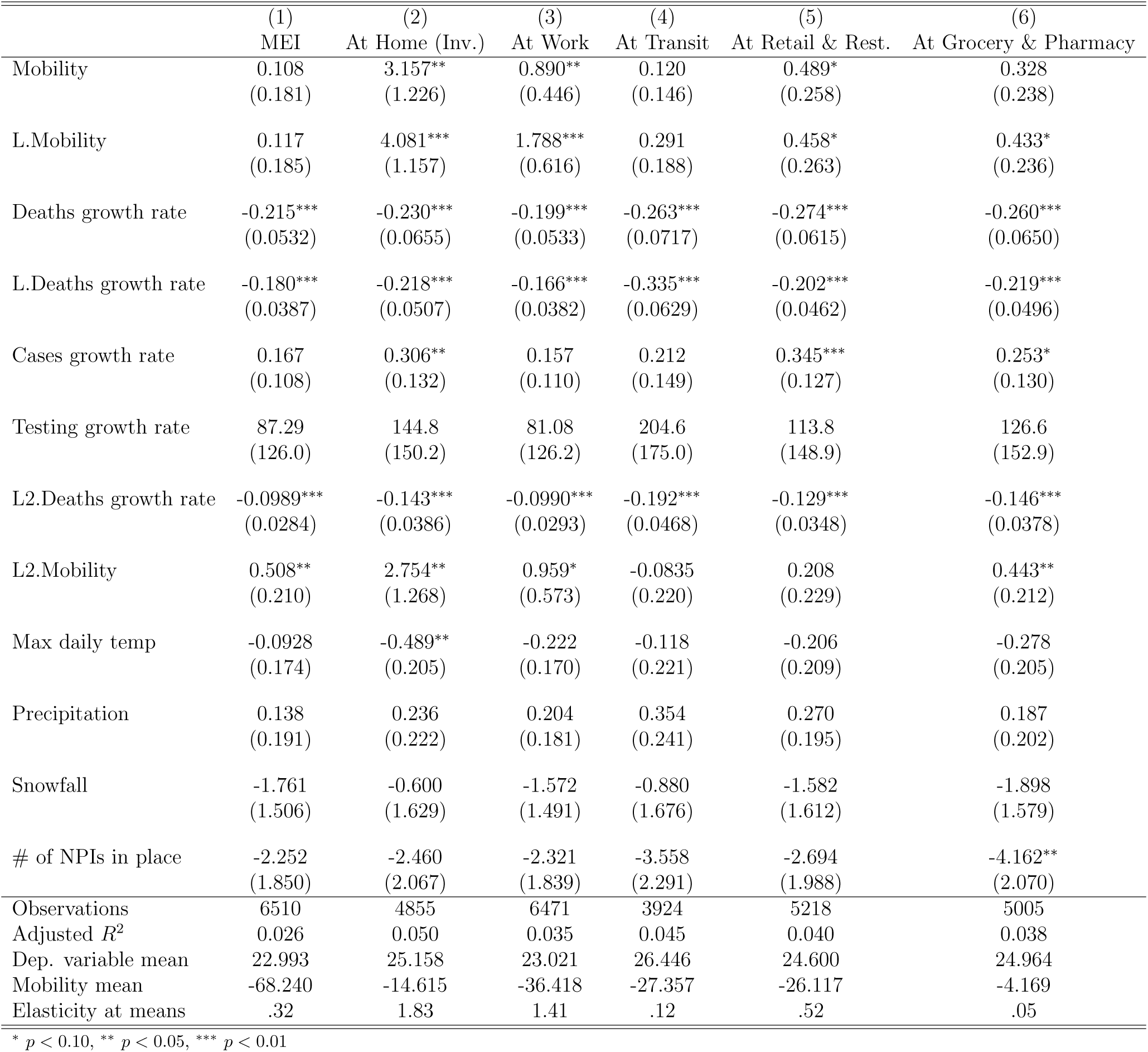
Estimates of Effect on 10-Week Ahead Deaths Growth Controlling for Non-Pharmaceutical Interventions

Controlling for Non-Pharmaceutical Interventions
|  | (1) | (2) | (3) | (4) | (5) | (6) |
| --- | --- | --- | --- | --- | --- | --- |
|  | MEI | At Home (Inv.) | At Work | At Transit | At Retail & Rest. | At Grocery & Pharmacy |
| Mobility | 0.108<br>(0.181) | 3.157**<br>(1.226) | 0.890**<br>(0.446) | 0.120<br>(0.146) | 0.489*<br>(0.258) | 0.328<br>(0.238) |
| L.Mobility | 0.117<br>(0.185) | 4.081***<br>(1.157) | 1.788***<br>(0.616) | 0.291<br>(0.188) | 0.458*<br>(0.263) | 0.433*<br>(0.236) |
| Deaths growth rate | -0.215***<br>(0.0532) | -0.230***<br>(0.0655) | -0.199***<br>(0.0533) | -0.263***<br>(0.0717) | -0.274***<br>(0.0615) | -0.260***<br>(0.0650) |
| L.Deaths growth rate | -0.180***<br>(0.0387) | -0.218***<br>(0.0507) | -0.166***<br>(0.0382) | -0.335***<br>(0.0629) | -0.202***<br>(0.0462) | -0.219***<br>(0.0496) |
| Cases growth rate | 0.167<br>(0.108) | 0.306**<br>(0.132) | 0.157<br>(0.110) | 0.212<br>(0.149) | 0.345***<br>(0.127) | 0.253*<br>(0.130) |
| Testing growth rate | 87.29<br>(126.0) | 144.8<br>(150.2) | 81.08<br>(126.2) | 204.6<br>(175.0) | 113.8<br>(148.9) | 126.6<br>(152.9) |
| L2.Deaths growth rate | -0.0989***<br>(0.0284) | -0.143***<br>(0.0386) | -0.0990***<br>(0.0293) | -0.192***<br>(0.0468) | -0.129***<br>(0.0348) | -0.146***<br>(0.0378) |
| L2.Mobility | 0.508**<br>(0.210) | 2.754**<br>(1.268) | 0.959*<br>(0.573) | -0.0835<br>(0.220) | 0.208<br>(0.229) | 0.443**<br>(0.212) |
| Max daily temp | -0.0928<br>(0.174) | -0.489**<br>(0.205) | -0.222<br>(0.170) | -0.118<br>(0.221) | -0.206<br>(0.209) | -0.278<br>(0.205) |
| Precipitation | 0.138<br>(0.191) | 0.236<br>(0.222) | 0.204<br>(0.181) | 0.354<br>(0.241) | 0.270<br>(0.195) | 0.187<br>(0.202) |
| Snowfall | -1.761<br>(1.506) | -0.600<br>(1.629) | -1.572<br>(1.491) | -0.880<br>(1.676) | -1.582<br>(1.612) | -1.898<br>(1.579) |
| # of NPIs in place | -2.252<br>(1.850) | -2.460<br>(2.067) | -2.321<br>(1.839) | -3.558<br>(2.291) | -2.694<br>(1.988) | -4.162**<br>(2.070) |
| Observations | 6510 | 4855 | 6471 | 3924 | 5218 | 5005 |
| Adjusted $R^2$ | 0.026 | 0.050 | 0.035 | 0.045 | 0.040 | 0.038 |
| Dep. variable mean | 22.993 | 25.158 | 23.021 | 26.446 | 24.600 | 24.964 |
| Mobility mean | -68.240 | -14.615 | -36.418 | -27.357 | -26.117 | -4.169 |
| Elasticity at means | .32 | 1.83 | 1.41 | .12 | .52 | .05 |

\* $p < 0.10$ , \*\* $p < 0.05$ , \*\*\* $p < 0.01$

In sum, it appears that non-pharmaceutical interventions have some beneficial effects on future COVID-19 cases even holding mobility fixed, though their impact on future deaths is less clear.

### 5.3 Time-Varying Mobility Effects

Lastly, I assess whether the marginal effect of mobility changes of a given size on COVID-19 outcomes has changed over the course of the pandemic thus far. To assess how the key results of the paper have evolved over recent weeks, I re-estimate the mobility IRFs presented in Section 4.3 (based on equation 2) repeatedly using 10-week rolling samples. Specifically, I estimate the IRFs first using data on the independent variables for the 10-week period ending with week 16 of 2020 (the week ending April 21), then using the 10-week period ending week 17, and so on until the latest week of available data.

To summarize how the results have changed over recent weeks, I plot the 8-week-ahead mobility elasticity (and its confidence interval) for both cases growth and deaths growth against the end-of-sample week. The results are shown in Figure 11. I focus on the three broadest measures of mobility: time spent away from home, time spent at work, and the MEI. The panels on the left are for cases growth, while the panels on the right are for deaths growth. The marginal effect of mobility on cases has been fairly stable over time, with no clear trend. However, the marginal effect on deaths has steadily fallen. One possible explanation is that over time mobility has become less lethal, in a sense, because of improved medical practices such as early infection detection and identification of some effective treatments.

**Figure 11:**
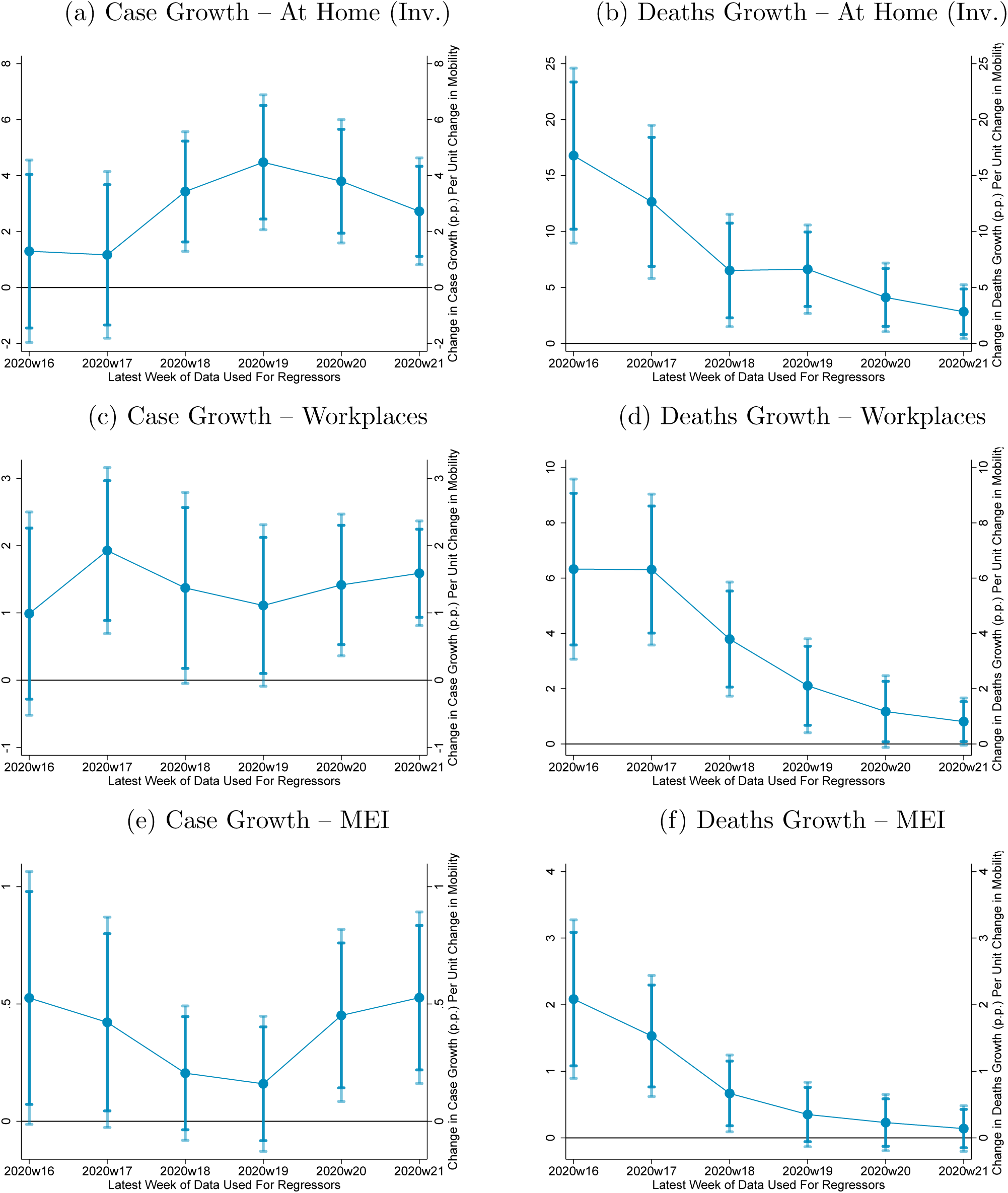
Rolling Regressions (10-Week Windows) Impact of Mobility on COVID-19 Outcomes 8-Weeks Ahead Note: Estimates of equation 2 in the text using panel local projections regressions. Shaded regions are 90% and 95% confidence intervals.

## 6 Conclusion

This paper sought to provide estimates of the full dynamic response of COVID-19 outcomes to exogenous movements in mobility. It uncovered several important findings. First, temperature is found to have a negative and significant effect on future COVID-19 cases and deaths. Second, controlling for weather, overall mobility is found to have a large positive effect on subsequent growth in COVID-19 cases and deaths. The effects become significant around 4 weeks ahead and persist through around 8 weeks for cases and 10 weeks for deaths. Third, the effects are found to be highly dependent on the local transmission rate of the virus: when the transmission rate is below one, mobility has small effects; when the transmission rate is above one, mobility has large effects. Fourth, the dynamic effects of mobility on cases were found to be generally similar across counties, but the effects on deaths were found to be higher for counties with older populations and counties with lower black or hispanic population shares. Lastly, I found that the marginal impact of mobility changes has been stable over recent weeks for cases, but it has ameliorated for deaths.

This is a first attempt at estimating the full dynamic impact of mobility and weather on COVID-19 outcomes. As noted in the beginning of the paper, this dynamic estimation is increasingly feasible because of the availability of high-frequency, real-time data along with sufficient passage of time since the initial outbreaks in most of the U.S.. I plan to regularly revisit the analyses in this paper as further days and weeks of data become available.

## Data Availability

All data used in the study were obtained from publicly available online sources, which are documented in the paper.

## Appendix A Supplemental Results

**Figure A1:**
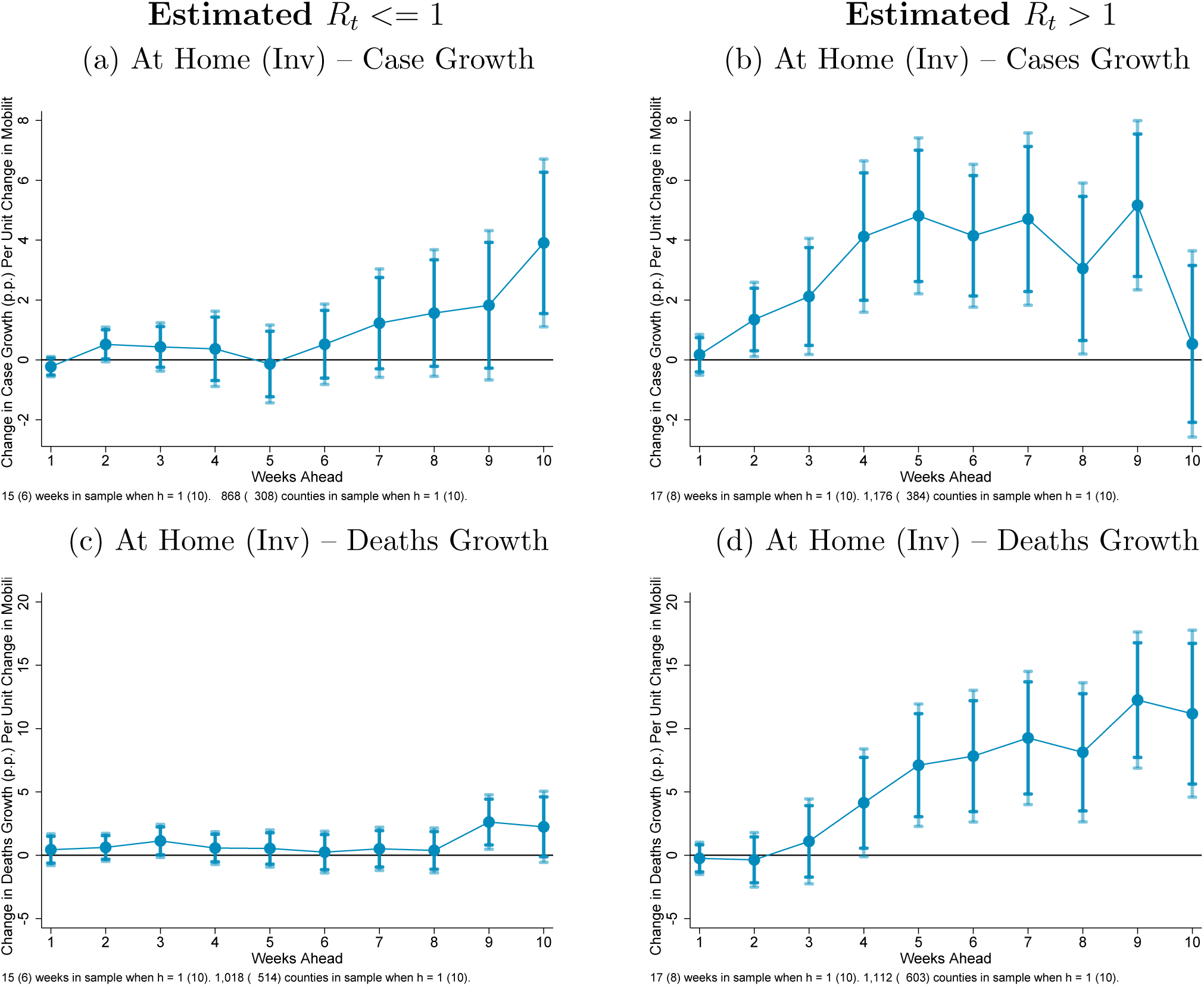
Dynamic Impacts of Mobility on Growth in COVID-19 Outcomes Note: Estimates of equation 2 in the text using panel local projections regressions. Shaded regions are 90% and 95% confidence intervals.

**Figure A2:**
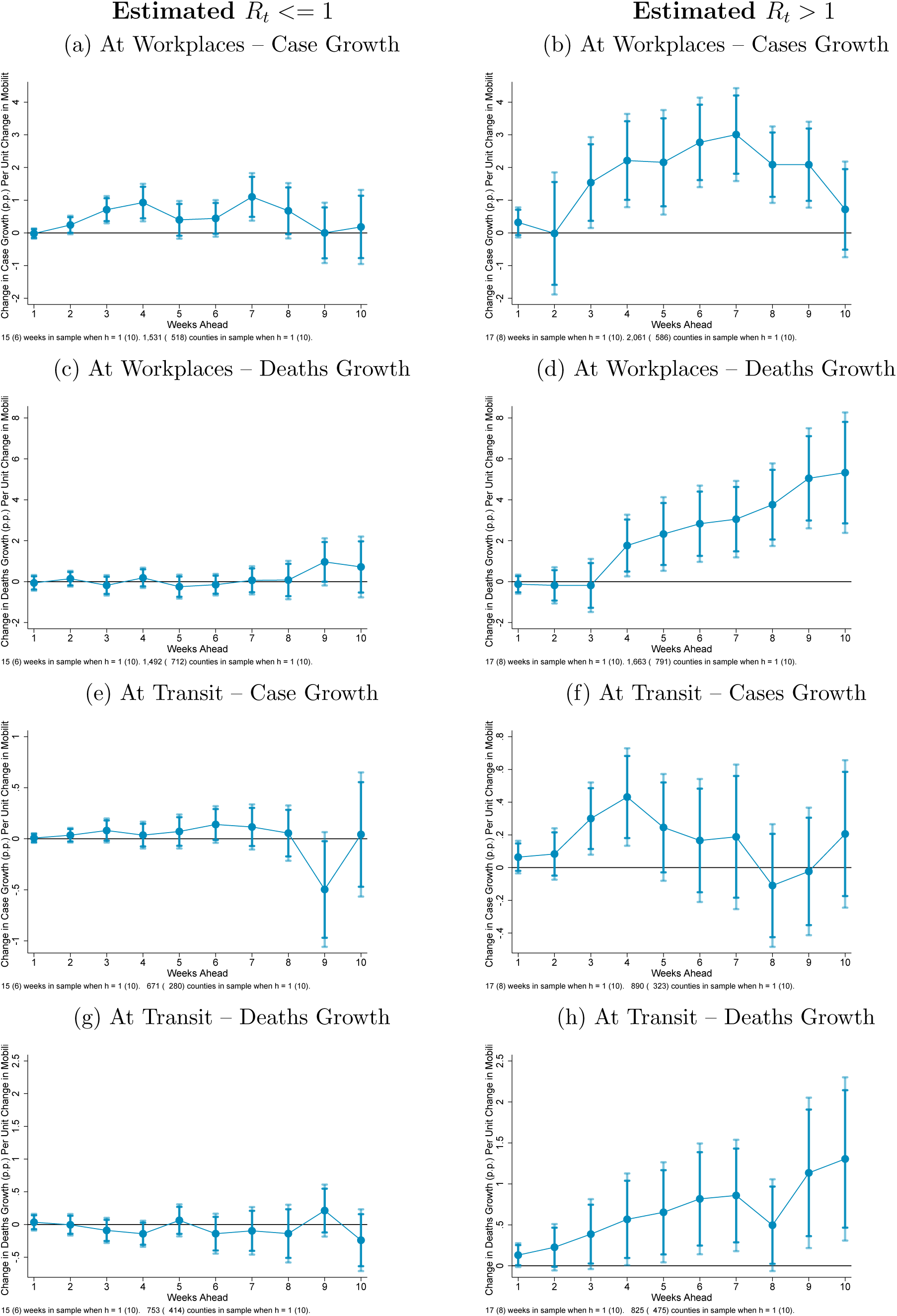
Dynamic Impacts of Mobility on Growth in COVID-19 Cases Note: Estimates of equation 2 in the text using panel local projections regressions. Shaded regions are 90% and 95% confidence intervals.

**Figure A3:**
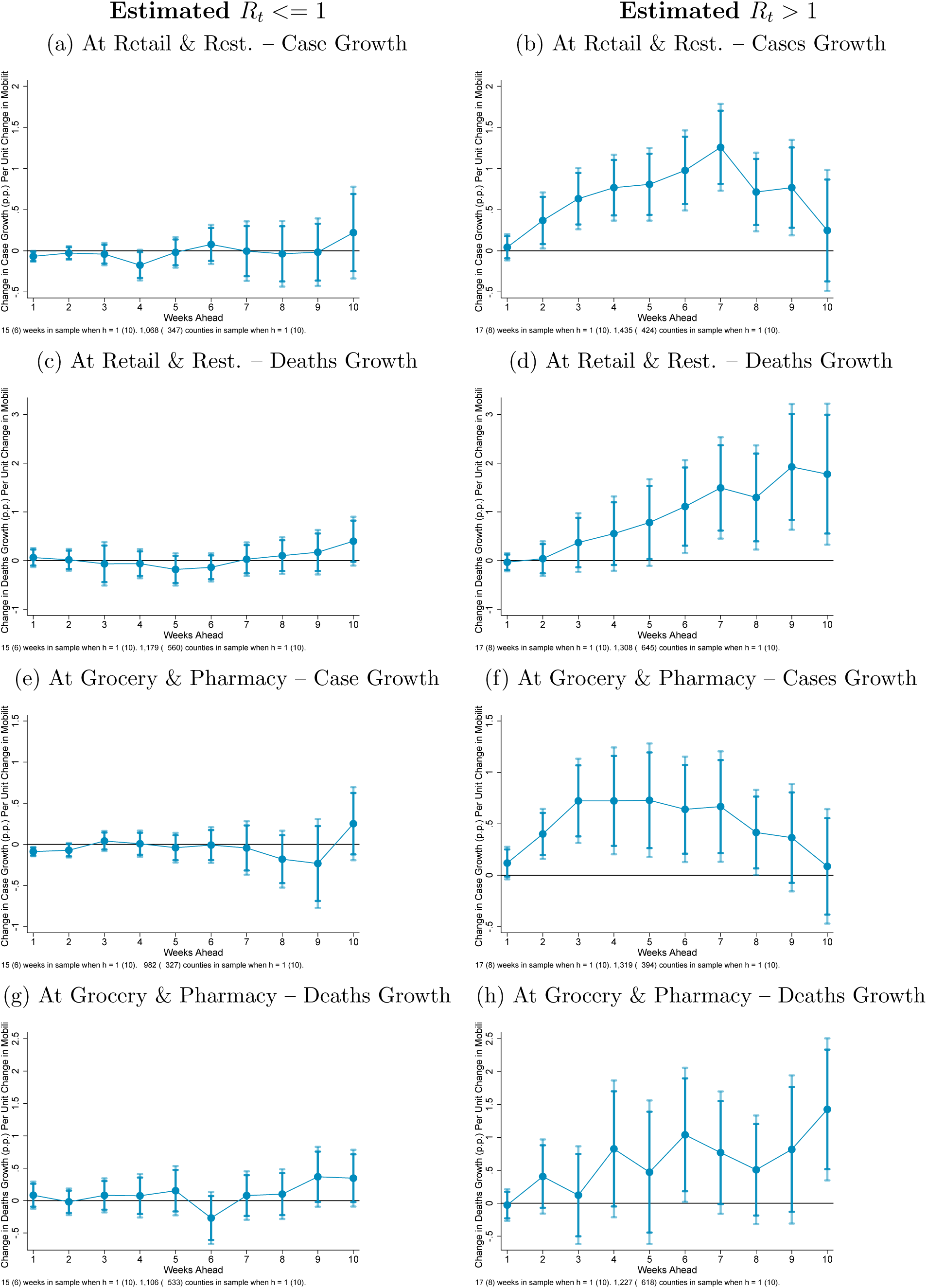
Dynamic Impacts of Mobility on Growth in COVID-19 Cases Note: Estimates of equation 2 in the text using panel local projections regressions. Shaded regions are 90% and 95% confidence intervals.

## Footnotes

1 For example, Chen and Spence (2020) document that across countries the drop in GDP in the first quarter of 2020 was highly correlated with declines in mobility.

2 May 13, 2020 speech at the Peterson Institute for International Economics.

3 See Nguyen, Gupta, Andersen, Bento, Simon, and Wing (2020) for a discussion of this literature as well as original evidence.

4 https://usafactsstatic.blob.core.windows.net/public/data/covid-19/covid_confirmed_usafacts.csv

5 There are two larges spikes in daily death counts in Figure 1. These are due to the addition by New York City and New Jersey of probable COVID-19 deaths based on retrospective reviews of deaths certificates. New York City added around 3,800 deaths on April 14 and New Jersey added 1,854 on June 27. I remove these added deaths from the data for the regression analyses in the paper. Their inclusion would yield large outliers in death growth rates which could unduly bias the results.

6 The data were accessed at https://www.google.com/covid19/mobility/.

7 The data were accessed at https://www.dallasfed.org/~/media/documents/research/mei/MEI_counties_scaled.csv.

8 https://raw.githubusercontent.com/Keystone-Strategy/covid19-intervention-data/master/complete_npis_inherited_policies.csv

## Notes

### Competing Interest Statement

The authors have declared no competing interest.

### Clinical Trial

Study is not a clinical trial.

### Funding Statement

No external funding.

### Author Declarations

No IRB approval required. Study does not use data on individual human subjects. Study uses county level aggregate data.

## References

Askitas, N., K. Tatsiramos, and B. Verheyden (2020): “Lockdown Strategies, Mobility Patterns and COVID-19,” Covid Economics, (23), 263–302.

Atkinson, T., J. Dolmas, C. Koch, E. F. Koenig, K. Mertens, A. Murphy, and K.-M. Yi (2020): “Mobility and Engagement Following the SARS-Cov-2 Outbreak,” FRB of Dallas Working Paper, (2014).

Badr, H., H. Du, M. Marshall, E. Dong, M. Squire, and L. M. Gardner (2020): “Social Distancing is Effective at Mitigating COVID-19 Transmission in the United States,” medRxiv.

Carleton, T., J. Cornetet, P. Huybers, K. Meng, and J. Proctor (2020): “Ultra-violet radiation decreases COVID-19 growth rates: Global causal estimates and seasonal implications,” Available at SSRN 3588601.

Chen, L., and M. Spence (2020): “Five lessons from tracking the global pandemic economy,” VoxEU.

Courtemanche, C., J. Garuccio, A. Le, J. Pinkston, and A. Yelowitz (2020): “Strong Social Distancing Measures In The United States Reduced The COVID-19 Growth Rate: Study evaluates the impact of social distancing measures on the growth rate of confirmed COVID-19 cases across the United States.,” Health Affairs, pp. 10–1377.

Desmet, K., and R. Wacziarg (2020): “Understanding Spatial Variation in COVID-19 across the United States,”.

Hsiang, S., D. Allen, S. Annan-Phan, K. Bell, I. Bolliger, T. Chong, H. Druckenmiller, L. Y. Huang, A. Hultgren, E. Krasovich, et al. (2020): “The effect of large-scale anti-contagion policies on the COVID-19 pandemic,” Nature, pp. 1–9.

Jamil, T., I. Alam, T. Gojobori, and C. M. Duarte (2020): “No evidence for temperature-dependence of the covid-19 epidemic,”.

Jordá, Ú. (2005): “Estimation and inference of impulse responses by local projections,” American economic review, 95(1), 161–182.

Kapoor, R., H. Rho, K. Sangha, B. Sharma, A. Shenoy, and G. Xu (2020): “God is in the Rain: The Impact of Rainfall-Induced Early Social Distancing on COVID-19 Outbreaks,” Available at SSRN 3605549.

Nguyen, T. D., S. Gupta, M. Andersen, A. Bento, K. I. Simon, and C. Wing (2020): “Impacts of state reopening policy on human mobility,” Discussion paper, National Bureau of Economic Research.

Soucy, J.-P. R., S. L. Sturrock, I. Berry, D. J. Westwood, N. Daneman, D. R. MacFadden, and K. A. Brown (2020): “Estimating effects of physical distancing on the COVID-19 pandemic using an urban mobility index,” medRxiv.

Unwin, H. J. T., S. Mishra, V. C. Bradley, A. Gandy, M. Vollmer, T. Mellan, H. Coupland, K. Ainslie, C. Whittaker, J. Ish-Horowicz, et al. (2020): “State-level tracking of COVID-19 in the United States,” Imperial College London, Unpublished Report.

Wilson, D. J. (2019): “Clearing the Fog: The Predictive Power of Weather for Employment Reports and their Asset Price Responses,” American Economic Review: Insights, 1(3), 373–88.

Xu, R., H. Rahmandad, M. Gupta, C. DiGennaro, N. Ghaffarzadegan, H. Amini, and M. S. Jalali (2020): “The Modest Impact of Weather and Air Pollution on COVID-19 Transmission,” medRxiv.

